# Covidization and decovidization of the scientific literature and scientific workforce

**DOI:** 10.1101/2024.09.13.24313660

**Authors:** John P. A. Ioannidis, Thomas A. Collins, Eran Bendavid, Jeroen Baas

**Affiliations:** Meta-Research Innovation Center at Stanford (METRICS), Stanford University, Stanford, CA, USA; Department of Medicine, Stanford University School of Medicine, Stanford, CA, USA; Department of Epidemiology and Population Health, Stanford University School of Medicine, Stanford, CA, USA; Elsevier, New York, NY, USA; Department of Health Policy, Stanford University School of Medicine, Stanford, CA, USA; Research Intelligence, Elsevier BV, Amsterdam, Netherlands

**Keywords:** COVID-19, citations, bibliometrics, authorship

## Abstract

We examined the growth trajectory and impact of COVID-19-related papers in the scientific literature until August 1, 2024 and how the scientific workforce was engaged in this work. Scopus indexed 718,660 COVID-19-related publications. As proportion of all indexed scientific publications, COVID-19-related publications peaked in September 2021 (4.7%) remained at 4.3-4.6% for another year and then gradually declined, but was still 1.9% in July 2024). COVID-19-related publications included 1,978,612 unique authors: 1,127,215 authors had ≥5 full papers in their career and 53,418 authors were in the top-2% of their scientific subfield based on a career-long composite citation indicator. Authors with >10%, >30% and >50% of their total career citations be to COVID-19-related publications were 376,942, 201,702, and 125,523, respectively. As of August 1, 2024, 65 of the top-100 most-cited papers published in 2020 were COVID-19-related, declining to 24/100, 19/100, 7/100, and 5/100 for the most-cited papers published in 2021, 2022, 2023, and 2024, respectively. Across 174 scientific subfields, 132 had ≥10% of their active influential (top-2% by composite citation indicator) authors publish something on COVID-19 during 2020-2024. Among the 300 authors with highest composite citation indicator specifically for their COVID-19-related publications, 41 were editors or journalists/columnists and another 23 had most of their COVID-19 citations to published items other than full papers (opinion pieces/letters/notes). COVID-19 massively engaged the scientific workforce in unprecedented ways. As the pandemic ended, there has been a sharp decline in the overall volume and high impact of newly published COVID-19-related publications.

**Significance statement:** COVID-19 massively mobilized the scientific workforce. Between 2020 and 2024, over 700,000 papers were published on COVID-19, including 2 million different authors. Across science, almost a third of authors at the top-2% of citation impact in their subfield published on COVID-19. There was a sharp decline in the proportion of COVID-19 papers across science after 2022 and an even more sharp decline in the proportion of COVID-19 papers reaching the highest level of citations. Authors with the highest COVID-19 citation impact prominently included many who were editors, journalists/columnists and opinion writers publishing massively. While other epidemics also witnessed sharp increases and subsequent decline in interest, the magnitude of the covidization and decovidization process is unique in the scientific literature to-date.

## INTRODUCTION

The COVID-19 pandemic was a major crisis that mobilized wide segments of the scientific workforce and led to a large corpus of scientific publications. Earlier analyses suggested that by August 1, 2021, more than 200,000 Scopus-indexed publications were related to COVID-19 (1). More than 700,000 unique authors were involved in these publications with contributions from authors specializing across all 174 subfields of science (according to the Science-Metrix classification) (1). Moreover, in 109 of the 174 subfields, at least 1 in 10 of the most-cited authors (according to a composite citation indicator) (2) had published something on COVID-19. Another analysis, focusing on 2020 and the first part of 2021 (3) found that almost all of the 100 most-cited publications across science in this timeframe were related to COVID-19. COVID-19-related work was on average more cited than work in other fields and this resulted in massive increases in the journal impact factor of some journals and the creation of a new generation of highly-cited scientists (3,4). A new citation elite of authors was created by those who massively boosted their citation metrics with their COVID-19 work (3). The massive growth and wide extent of COVID-19-related publications has led to discussions and concerns about the covidization of the research enterprise (5,6).

Since this earlier work, the pandemic has run its course and ended (7), but some of the repercussions continue to exist and attract scientific interest. The current analysis aims to assess the growth trajectory and impact of COVID-19-related papers in the scientific literature during a period of 4.5 years (from the beginning of 2020 to 1 August 2024) and to examine how the scientific workforce was engaged in this work and potentially disengaged after the crisis had de-escalated. One might hypothesize that both volume and impact may have peaked and declined, perhaps with some time lag with the course of the hard pandemic indicators (e.g. COVID-19-related deaths). However, it is unclear whether the pattern of decline is qualitatively different to the decline seen for other acute epidemics, such as Zika, H1N1, or Ebola that also attracted sudden scientific attention in the past, albeit at much smaller scale (8–15). Hence, we are using Scopus data until August 2024 to understand better the process of covidization and the potential pace of a respective decovidization of the scientific literature.

## METHODS

### Protocol

The protocol for this meta-research analysis was registered in advance at Open Science Framework: https://osf.io/84bdt.

### Search strategy for eligible COVID-19 publications

We used a copy of the Scopus database (16) extracted on 1 August 2024. We followed the exact same search strategy for identifying COVID-19 publications, as in previous work (1), using the query: TITLE-ABS-KEY(sars-cov-2 OR ‘coronavirus 2’ OR ‘corona virus 2’ OR covid-19 OR {novel coronavirus} OR {novel corona virus} OR 2019-ncov OR covid OR covid19 OR ncovid-19 OR ‘coronavirus disease 2019’ OR ‘corona virus disease 2019’ OR corona-19 OR SARS-nCoV OR ncov-2019) AND PUBYEAR > 2018. As in that previous work, we also further filtered the dataset using the Elsevier International Center for the Study of Research Lab infrastructure to publications indexed (loaded) in Scopus in 2020 or later, and with the publication year of 2020 or greater. We evaluated publication dates by month, where month was available. When publication month was either unavailable or exceeded the indexing date, we used the indexing date instead. This accounts for cases where an article is published early, but the official journal issue appears later. We were interested in the date at which publications became available to the public rather than official publication dates.

Also similar to previous work (1), we considered both publications in peer-reviewed venues and preprints. To avoid double counting of the same item published both in a peer-reviewed journal and as a preprint, or/and in two preprint servers, we identified and filtered out duplicates by matching against author names and titles. We applied methods used for unstructured reference linking (17) ranking documents based on similarity of fields. The best match is based on the overlap between words in the title and author names. We excluded preprints that link to either a non-preprint item, such as journal articles, or an earlier preprint. The result of this step is the exclusion of 38,641 preprints.

We further focused on the 6,653,434 “active” authors who have at least one Scopus-indexed publication since 2020 and who have also authored in their entire career at least five Scopus-indexed papers classified as articles, reviews or conference papers (“full articles”). This excludes authors with limited recent presence in the scientific literature and some author IDs that may represent split fragments of the publication record of more prolific authors.

### Field classification

All authors were assigned to their most common field and subfield discipline of their career by field breakdown of their publications. Similar to previous work (1), we used the Science Metrix classification of science, which is a standard mapping of all science into 21 main fields and 174 subfield disciplines (18).

### Influential scientists

We also examined how COVID-19 has affected the publication portfolio of researchers whose work has the largest citation impact in the literature. We used the career-long citation statistics calculated with the Scopus database of 1 August 2024 (19). Each author has been assigned to the main field and main subfield based on the largest proportion of publications across fields, and analysis is restricted to the top 2% authors per Science Metrix subfield. We have previously developed a composite citation indicator (2,20,21) that combines information on six indices: total citations, Hirsch h-index, Schreiber hm-index, citations to single-authored papers, citations to first- or single-authored papers, and citations to first-, single- or last-authored papers. Self-citations are excluded from all calculations. Accordingly 212,343 scientists can be classified as being in the top 2% of their main subfield discipline based on the citations that their work received through the end of 2023. Of those, 174,764 were active and had published at least one paper also in 2020-2024.

### Authors with high citation impact of their COVID-19 publication record

We also identified the 300 authors whose COVID-19 publications to date had the highest citation impact. The same composite citation indicator for all authors was calculated limited to COVID-19-related publications and the top-300 highest scoring authors were selected.

### Analyses

The following descriptive analyses were done on the data generated as stated above. First, we obtained the total number of COVID-19-related publications and the total number of authors involved (overall; among those who published at least one item in 2020-2024 and had at least 5 full papers in their career; and among those in the top-2% of impact who also published at least one item in 2020-2024). We also noted how many authors in these three authors’ sets had >10%, >30% and >50% of their total career-long citations be to COVID-19-related publications.

Second, we evaluated the monthly publication of COVID-19-related items from January 2020 until mid-2024 to identify if there was a peak and if so, how rapid and extensive the decline had been afterwards. We used both absolute counts of published items as well as proportions of total Scopus items. Proportionate analysis is more reliable to assess changes over time, since the number of total published items indexed each month is increasing over time and indexing may have some time lag thus absolute counts may be lower in very recent months. The curve of monthly COVID-19-related items was also juxtaposed against the bursts of bibliographic coverage of other epidemics that attracted sudden attention in the past. For these analyses, published items were found in Scopus using the search terms “Zika”, “H1N1”, and “Ebola” with the period of interest starting in 2016, 2009, and 2014, respectively. Given the older time frames where monthly data may not be accurate, we used annual data for the other epidemics. Furthermore, in exploratory analyses we assessed the citation of COVID-19-related papers overall and per month versus other papers published in 2020-2024.

Third, we evaluated how many of the top-100 most-cited papers published in 2020 were on COVID-19, and how this declined for the 100 most-cited papers published in 2021, 2022, 2023, and the first part of 2024. For generating the lists of the top-cited papers, we used citations received as of 1 August 2024. In order to see if the most-cited COVID-19 highly-cited papers were gradually falling off the top-cited list of the most-cited items across all science, we compared these numbers against figures on the top-100 of each year based on citations received until the end of 2020, end of 2021, end of 2022, and end of 2023.

Fourth, we evaluated for each of the 174 scientific subfields what was the proportion of active (those with at least one published item in 2020-2024) top-2% influential scientists who had published something on COVID-19 in 2020, in 2021, in 2022, in 2023, and in the first part of 2024. We then noted how many subfields in each year had at least 5%, 10%, 30%, and 50% of their active top-2% influential scientists publishing anything on COVID-19. We hypothesized that after an initial mobilization of expertise across many diverse fields, over time only specific fields would continue to have many top-cited scientists publishing on COVID-19.

Fifth, for each of the 300 authors with the highest composite citation indicator for their COVID-19 publications, we recorded how many citations their COVID-19-related work had received in 2020, in 2021, in 2022, in 2023, and in first part of 2024. We then examined whether the median proportion of citations received decreased over time. We also examined to what extent the list of the 300 top-cited authors for COVID-19 related work by 1 August 2024 overlapped with the list of the 300 top-cited authors for COVID-19-related work by 1 August 2021 generated in our previous analysis (1). This allowed to examine if the COVID-19 citation elite was largely determined in the early phases of the pandemic. Upon collating the list of the 300 top-cited authors, we observed that many, especially those at the very top of the list, were editors or journalists/columnists rather than scientists contributing original work. Therefore, we also assessed how many of the 300 were editors or journalists/columnists with most of all COVID-19 citation impact related to that role; and how many were other scientists who had >50% of their citations to COVID-19-related publications given to items that were not full papers (“articles”, “reviews” or “conference papers” in the Scopus classification of published items), i.e. largely to opinion pieces, letters and shorter notes.

## RESULTS

### COVID-19 published items and authors

As of 1 August 2024, Scopus classified 718,660 published items as relevant to COVID-19. This number accounts for 3.6% of the 19,971,245 published items across all science in the period 1 January 2020 until 1 August 2024. The COVID-19-related published items were classified by Scopus as articles (453,331, 63.1%), reviews (59,920, 8.3%), letters (43,781, 6.1%), conference papers (42,354, 5.9%), non-matched preprints (33,166, 4.6%), notes (25,423, 3.5%), editorials (23,486, 3.3%) and other items (37,199, 5.2%).

The 718,660 COVID-19-related items include 1,978,612 unique authors (with different Scopus IDs), amounting to 9.7% of the 20,365,211 author IDs who have published at least 1 paper of any type and on any topic in 2020-first part of 2024. Among the 6,653,434 authors who have published anything that is Scopus-indexed in 2020-first part of 2024 and who have also authored in their entire career at least five Scopus-indexed full papers (classified as articles, reviews or conference papers), 1,127,215 of these authors (16.9%) had at least one published and indexed COVID-19-related item. When further limited to the top-2% in citation impact, there were 53,418 authors out of 174,764 (30.6%) with at least one published and indexed COVID-19 item.

A total of 921,211 authors (376,886 with at least 5 full papers, 2,471 among the top-2%) had >10% of their total career-long citations be to COVID-19-related work. This represented 4.5%, 5.7%, and 1.4%, of the total authors across science in these categories, reprctively. The respective number of authors with >30% of their total career-long citations be to COVID-19-related work was 703,319 (201,707 with at least 5 full papers, 602 among the top-2%). With a >50% citation threshold, the number of authors was 584,397 (125,665 with at least 5 full papers, 210 among the top-2%).

### Monthly COVID-19-related publications

Figure 1 shows the monthly COVID-19-related publications in absolute numbers (Figure 1A) and as percentage of total published publications across all science (Figure 1B). As shown, the peak percentage was reached in September 2021 with 4.7% of all items published that month and the percentage remained very high, at 4.3-4.6%, for another year. This period corresponds also to last peak of COVID-19-related deaths globally (February 2022). There was steady decline afterwards, but even in July 2024 COVID-19-related papers still accounted for 1.9% of all published items across science. This represented a 60% decline from the peak value. Conversely, COVID-19 deaths recorded globally had declined by more than 95% of the last peak (February 2022) by summer 2023 and remained as low or lower after this.

**Figure 1.**
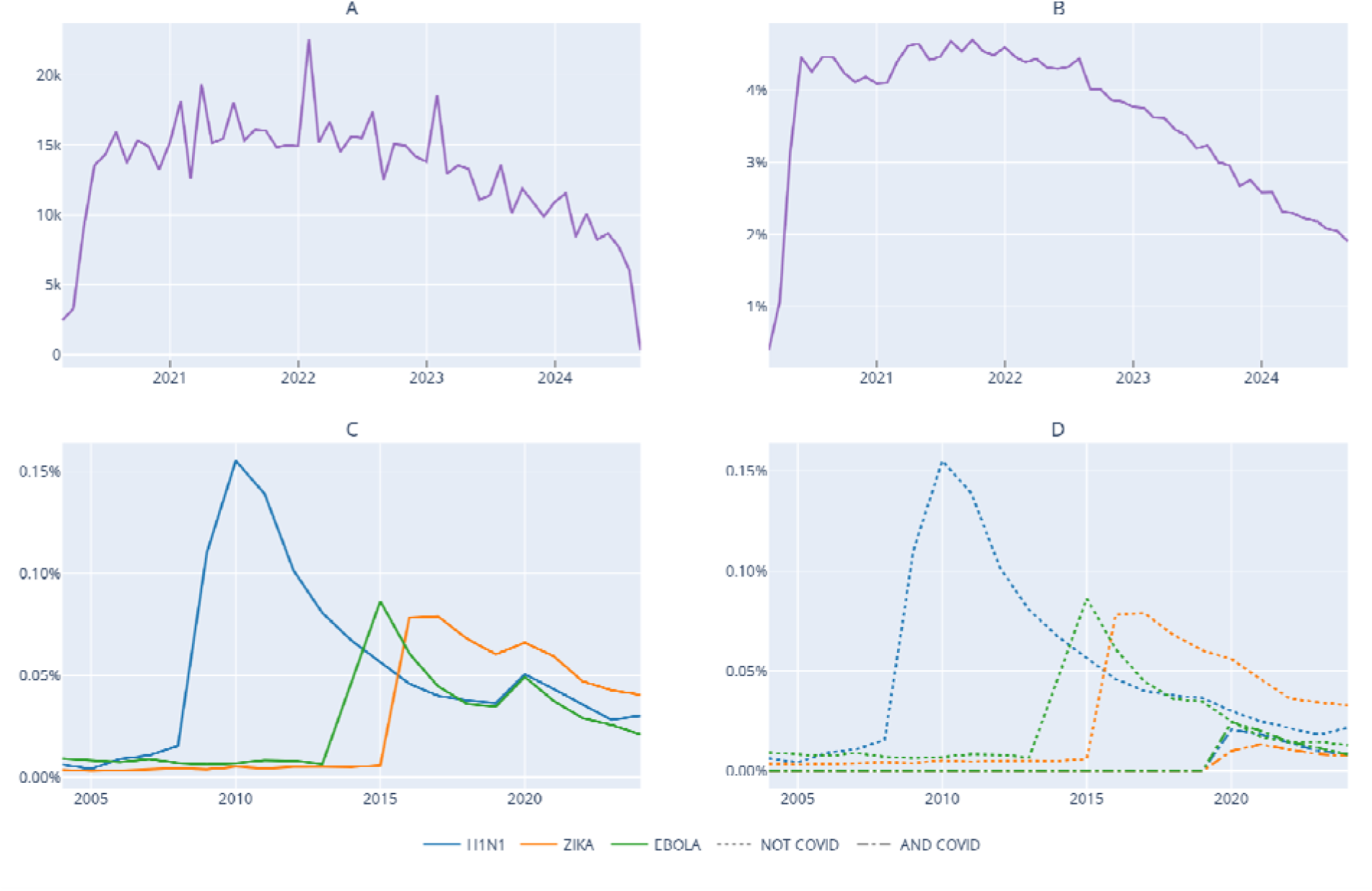
A. Monthly COVID-19 related publications in 2020-first part of 2024. B. Percentage of COVID-19-related publications in 2020-first part of 2024 among all science publications. C. Percentage of annual Zika, H1N1, and Ebola-related publications among all science publications. D. Percentage of annual Zika, H1N1, and Ebola-related publications among all science publications, separating those that are also COVID-19-related.

In the cases of Zika, H1N1, and Ebola, there was also a prompt steeper decline after the peak (Figure 1C). Of interest, with the emergence of the COVID-19 pandemic in 2020, these other epidemic infections also showed a renewal of interest with an increase of their relative percentage during 2020. The increase was entirely due to papers that were also COVID-19-related (Figure 1D).

The COVID-19-related items have received 10,453,962 citations in total. As shown in Figure 2A, monthly citations have remained very high even in the first part of 2024, but they do not seem to increase further and they are distributed to an increasing volume of total COVID-19-related papers. Among all papers across science published in 2020 forward, COVID-19-related papers reached a peak 37% of total citations by April 2020, but had steeply declined to 3% by mid-2024 (Figure 2B). As shown in Figure 3, COVID-19-related papers were far more cited than other papers in 2020 and in 2021 and this resulted even in a shift of the citation distribution across all science (Figure 3A and B). This shift pattern was no longer discernible in 2022 and 2023 (Figure 3C and D).

**Figure 2.**
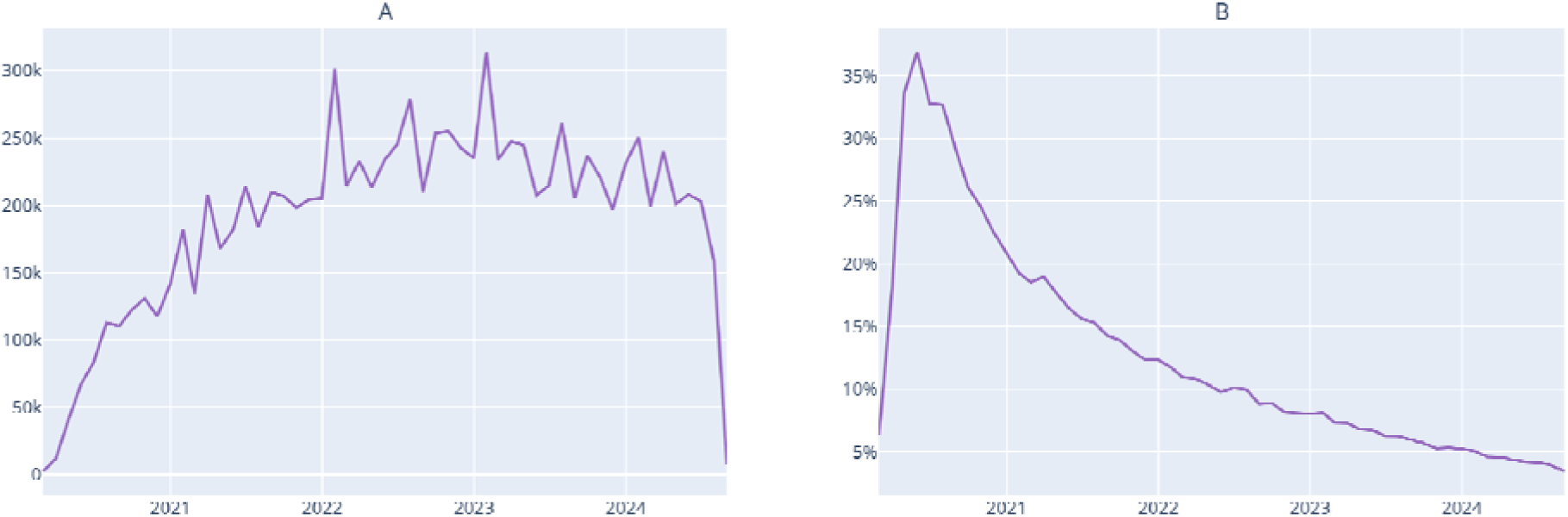
A. Monthly citations to COVID-19 related publications in 2020-first part of 2024. B. Percentage of citations to COVID-19 related publications as a proportion of citations given that month to all Scopus publications published in 2020-first part of 2024.

**Figure 3.**
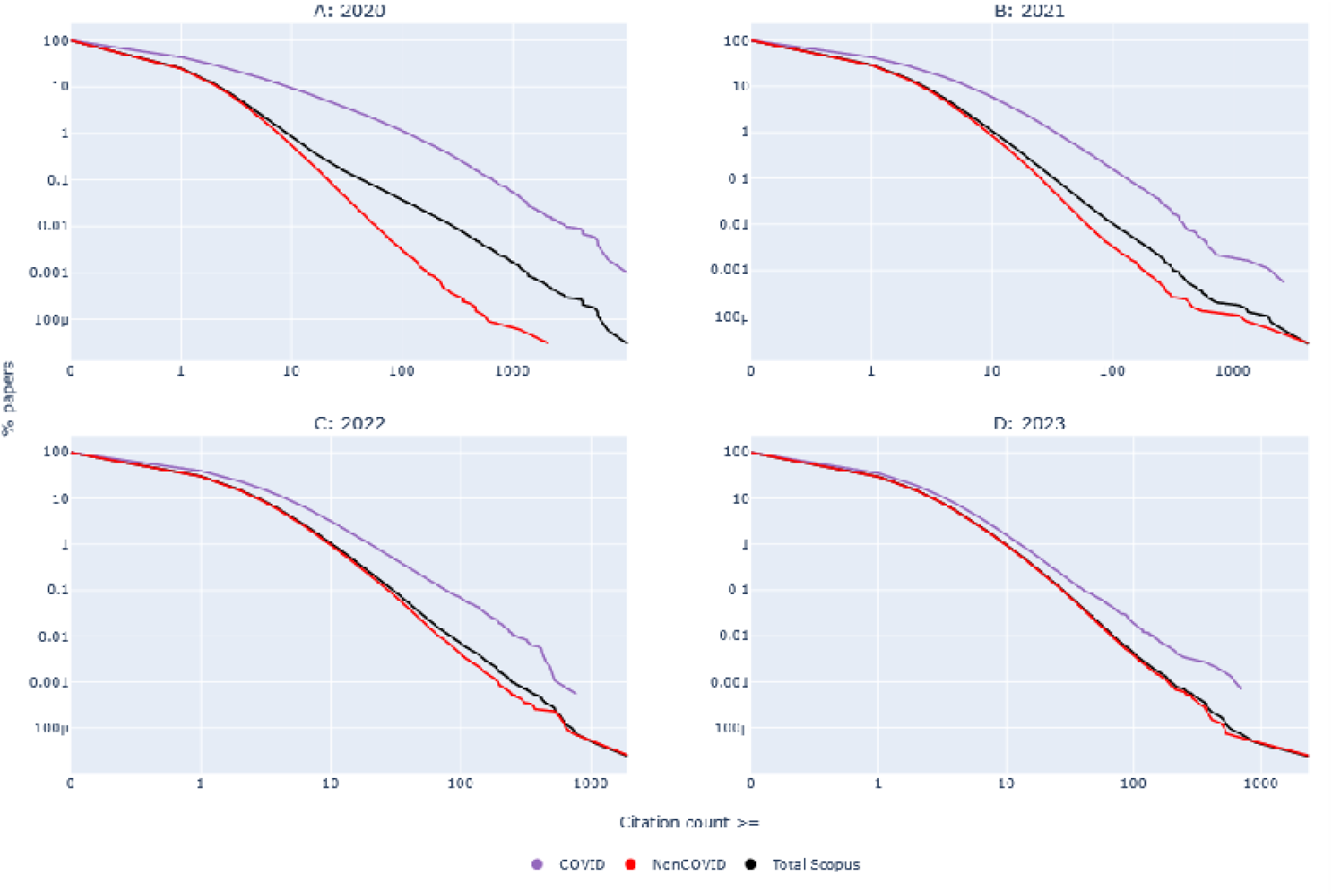
Distribution of citations received during the year of publication to papers published in 2020, 2021, 2022, and 2023 according to whether they were COVID-19-related, not COVID-19-related and overall (all science).

### COVID-19-related items among the top-100 most-cited across science

As shown in Table 1, by mid-2024, COVID-19-related items accounted for the large majority of the top-100 most-cited items across science published in 2020, but the proportion sharply declined afterwards and for the first part of 2024 only 5 of the top-100 most-cited items were related to COVID-19. The proportion of COVID-19-related items among the top-100 most-cited items across science was the highest in 2020 and 2021 and then sharply declined in subsequent years.

**Table 1.** Percentage of COVID-19-related items among the top-100 most-cited items across all science.

Number of COVID-19 items among top-100 most-cited Citation analysis across all science performed at
| Year of publication | End-2020 | End-2021 | End-2022 | End-2023 | 2024 (first part) |
| --- | --- | --- | --- | --- | --- |
| 2020 | 92 | 92 | 84 | 69 | 65 |
| 2021 | N/A | 57 | 47 | 30 | 24 |
| 2022 | N/A | N/A | 47 | 29 | 19 |
| 2023 | N/A | N/A | N/A | 4 | 7 |
| 2024 (first part) | N/A | N/A | N/A | N/A | 5 |

N/A: not applicable

### Involvement of 2% top-cited scientists in COVID-19-related publications across scientific subfields

As shown in Table 2, in every calendar year the large majority of the 174 subfields of science had at least 5% of their top-2% most-cited authors published something on COVID-19 and the majority of fields had even >10% of their top-cited authors publish on COVID-19. In a modest minority of subfields, every year >50% of the top-cited authors published on COVID-19, but this was limited by 2023 to only Emergency and Critical Medicine, Virology, Public Health, and Geriatrics (2024 calendar year data are not yet complete). When considering the entire 2020-first part of 2024 period, almost all scientific subfields (157/174, 90%) had at least 5% of their top-cited authors publish on COVID-19, and 17 scientific subfields (10%) had more than half of their top-cited authors publish on COVID-19.

**Table 2.** Scientific subfields (among a total of 174) that had at least 5%, 10%, 30% or 50% of their 2% top-cited authors (based on composite citation indicator, whole career impact to end-2023) publish at least one COVID-19-related item in each calendar year and overall in 2020-2024 (until 1 August 2024)

|  | Subfields with representation among 2% top-cited authors |  |  |  |
| --- | --- | --- | --- | --- |
|  | >=5% | >=10% | >=30% | >=50% |
| Year |  |  |  |  |
| 2020 | 120 | 93 | 22 | 2 |
| 2021 | 138 | 107 | 38 | 12 |
| 2022 | 134 | 109 | 40 | 9 |
| 2023 | 129 | 100 | 35 | 4 |
| 2024 (first part) | 109 | 81 | 15 | 2 |
| 2020 to first part of 2024 | 157 | 132 | 71 | 17 |

### Authors with highest citation impact for COVID-19 publications

Supplementary Table S1 shows the 300 authors with the highest impact on their COVID-19-related publications based on a composite citation indicator limited to their COVID-19-related items and excluding self-citations. Among these 300 authors, the median proportion of the citations received to their COVID-19 work was 35% in 2020, 59% in 2021, 62% in 2022, 54% in 2023, and 45% in the first half of 2024 suggesting that for most of them in each calendar year COVID-19 was their main source of citation impact with only modest decline after a peak in 2022. 41 of these 300 authors were editors of journalists/columnists in major journals and another 23 had received >50% of their citations to COVID-10-related publications to items that were not full papers (articles/reviews/conference papers), but typically opinion pieces (editorials and commentaries), letters/correspondence, or short notes, most of them published very early in the pandemic. Table 3 shows the 40 non-editor/journalist/columnist authors with highest composite citation indicators of COVID-19-related work, revealing a large diversity of main subfields of work and a scatter across very diverse countries and institutions. 33/40 had received >30% of their total citations in 2020-first part of 2024 for COVID-19-related publications. Of the 300 most-cited authors for their COVID-19-related work as of 1 August 2021, 186 had remained on the top-300 list when data to 1 August 2024 were considered.

**Table 3.**
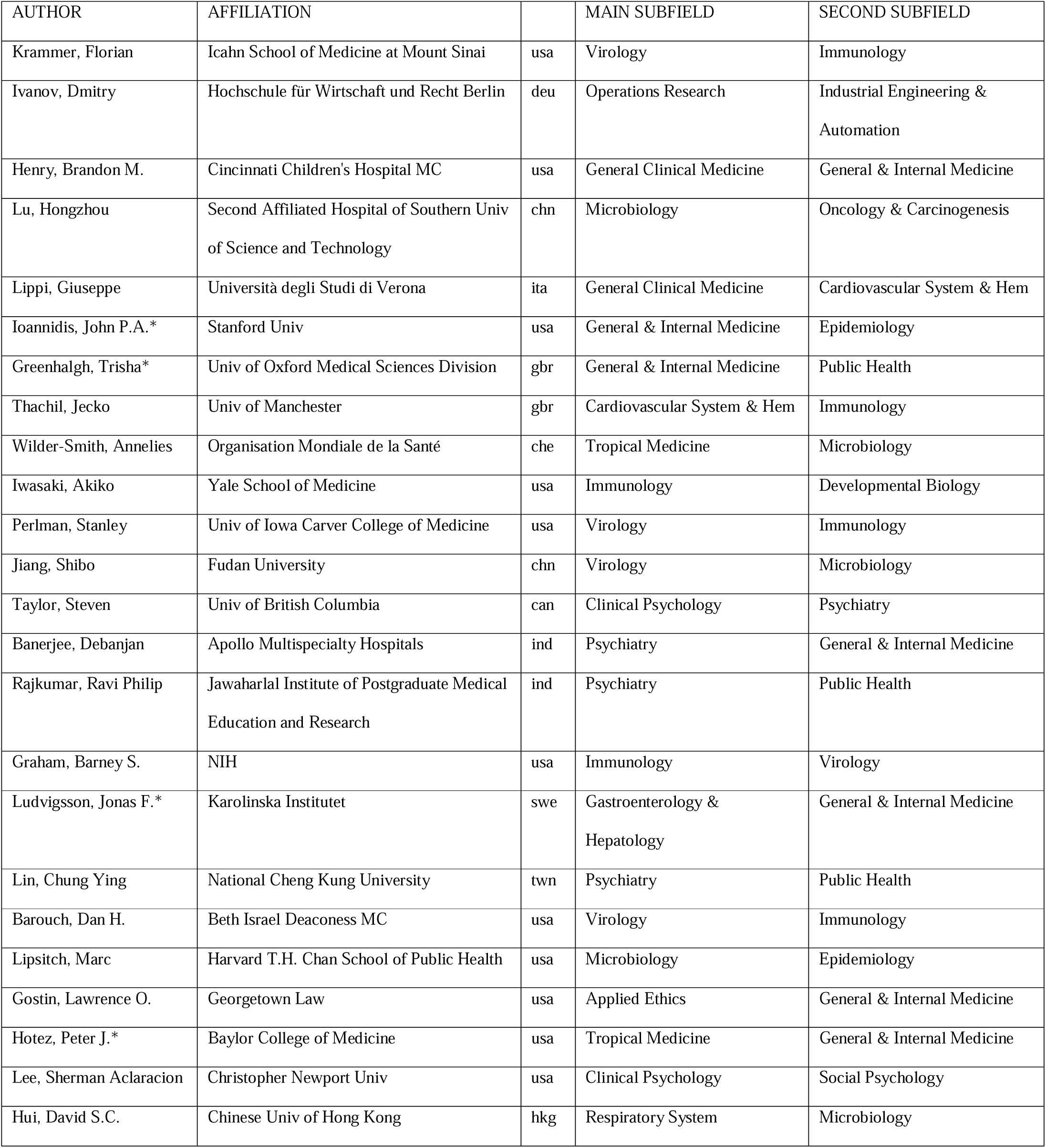
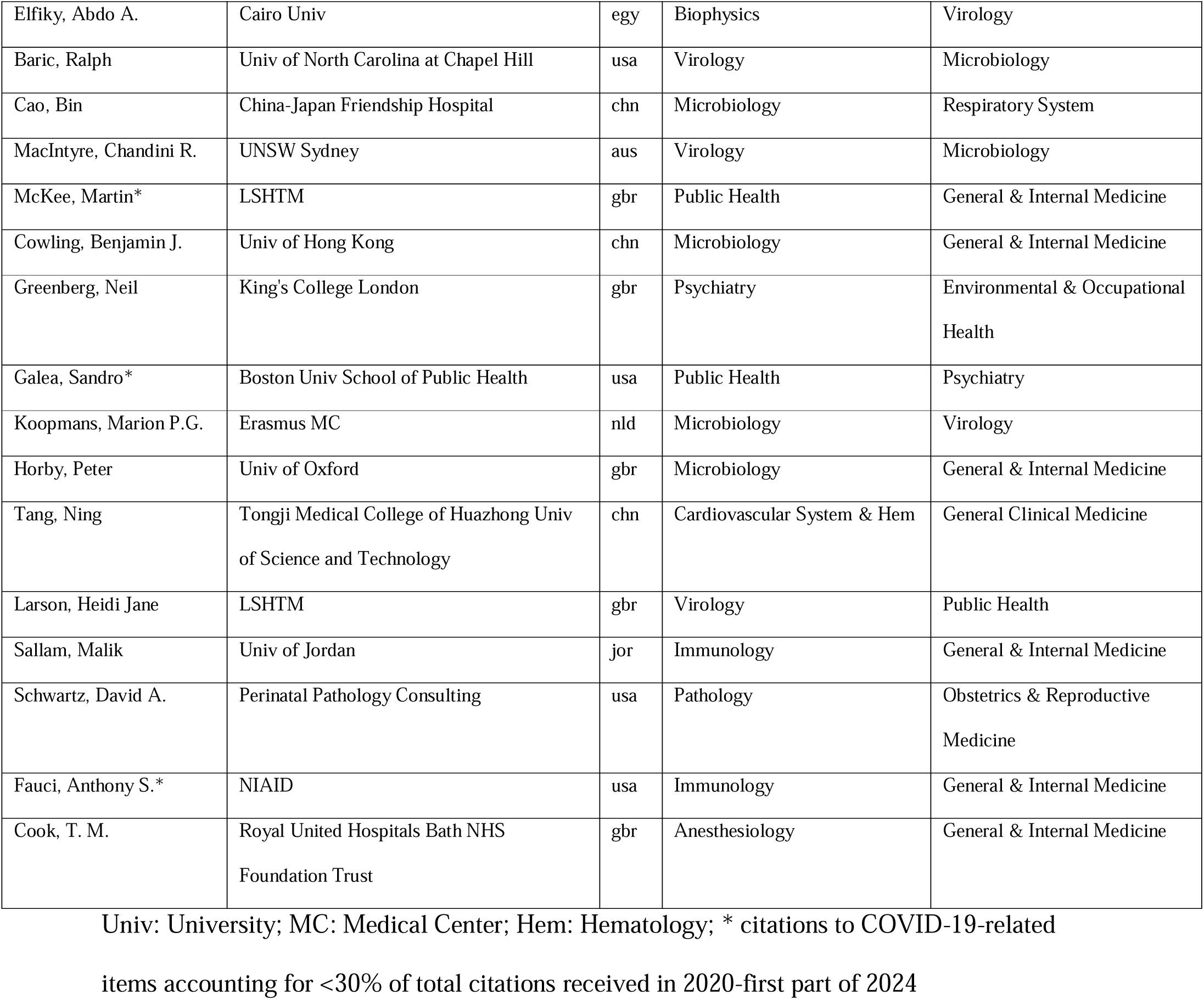
Top-40 authors with highest composite citation indicator for COVID-19-related published items during 2020-First Part of 2024. Editors and journalists/columnists are excluded.

## DISCUSSION

This meta-research analysis shows that over 700,000 COVID-19-related published papers were indexed in Scopus in 2020-first part of 2024 and this work involved 2 million different authors. This amounted to approximately 4% of the scientific literature indexed in Scopus and approximately 10% of the publishing scientific workforce. The proportion of authors who were involved in some COVID-19 publications was even higher among scientists with a track record of at least 5 published items (17%) and even more so among the top-2% most cited scientists (31%), presumably because when authors are more prolific they are more likely to have been involved in at least one COVID-19-related paper among many others. Nevertheless, COVID-19 also created a new large scientific workforce that has been highly committed to it: for more than half a million authors, more than half of their citations to-date are to COVID-19-related items. COVID-19-related papers practically monopolized the top-ranks of the most-cited publications across all science during 2020 and 2021. Since then, there has been a sharp decline in the number of extremely highly cited COVID-19 papers. There has been also a sharp decline in the continuing volume of COVID-19 publications after the fall of 2022. This is consistent with other prior epidemics where publication volume quickly declined after their acute phase. However, even by mid-2024 very large numbers of COVID-19 papers continue to be published, accounting for 1.9% of all scientific papers, while at the same time COVID-19 deaths currently represent approximately a log10 scale lower proportion of all global deaths. COVID-19-related papers have already received more than 10 million citations, but the rate of new citations has stabilized and it is spread more thinly across a much larger volume of existing COVID-19-related papers. In the large majority of the 174 subfields of science, there was still a sizeable number of top-cited authors who have continued to published COVID-19-related work even during 2023 and the first part of 2024. However, only 4 subfields have most of their top-cited authors continue to publish on COVID-19 by 2023. The top-cited authors within the COVID-19 literature cover a diverse range of disciplines and most of them yield the majority of their citations through COVID-19-related work.

The overall picture suggests a massive covidization and rapid decovidization of the scientific literature. The magnitude of these processes is unprecedented: the monthly volume of COVID-19 published papers at the covidization peak exceeded the volume of papers published by entire classic large disciplines such as Neuroscience, Ecology, Psychology, or Agriculture and it was more than an order of magnitude larger than the literature in fields like Mathematics (21). Even with ∼60% decline since then, the number of monthly papers has remained very large in absolute numbers even in mid-2024. Some papers may represent work that was done a while ago and took time to get published. Looking forward, funding and incentives may regulate interest in future COVID-19 publications. Some journal editors in pursuit of boosting their impact factors may still have a perception that COVID-19 papers are more cited than other papers (4,22,23), although the advantage seems to have decreased or even disappeared currently. Teams that shifted their interest to COVID-19 may not find it easy to disengage and move back to their previous research agendas. Moreover, a large new scientific workforce has been created, counting more than half a million authors whose track record to-date is mostly on COVID-19. Even in the absence of a new resurgence of a coronavirus pandemic, large numbers of COVID-19-related publications may continue to be released for the foreseeable future. The lesser citation impact that they enjoy may nevertheless gradually create an increasing disincentive. Further follow-up of the trajectory of the COVID-19 corpus will be interesting to pursue to understand better its complex dynamics.

The involvement of scientists in COVID-19 research was previously shown (1) to cover all 174 subfields of science already by the end of 2020. Wide involvement of very diverse fields has continued until mid-2024. Interdisciplinarity is desirable. Conversely, the occupation of experts with research that is remote from their main expertise may be problematic. There is a large number of studies suggesting that much COVID-19-related research has been of low quality (24–30). It is unknown whether a more voluminous literature may be associated with better quality currently and in the future. By 2023, only very few scientific subfields (all of them highly relevant to COVID-19) had the majority of their top-cited authors involved in COVID-19-related publications. This may herald some more focused, relevant expertise involved in currently published work.

Finally, COVID-19 not only generated a large new workforce committed to its study, but also generated a new citation elite (3). Our current analysis show that the highest citation impact scientists are usually highly focused on COVID-19 work, and this has continued for 4.5 years already with relatively little change overall. There is also a minority of influential COVID-19 researchers, nevertheless, who are also heavily cited in other scientific areas as well. A large number of the most influential authors on COVID-19 have been editors or journalists and columnists, mostly in high impact journals such as Nature, Science, BMJ, Lancet, and JAMA. Many others have obtained most of their citations to COVID-19 work through opinion pieces and letters/correspondence items rather than full articles. Many of these pieces were published very early in the pandemic and even though they were opinions or had limited data they attracted tremendous citations as they were available to be cited by the large volume of papers that followed. It has been observed across diverse scientific fields that early papers that achieve highly-cited status become ossified as references and new work, even if better, may not replace them (31). Moreover, the composite citation indicator that we used gives substantial weight to single- or first-authored publications. This may partly explain why several editorialists and columnists rank so highly. However, this alone would not suffice to rank them in the top positions of impact. The main reason is that many of these people authored extreme numbers of editorial or journalistic articles, some of which attracted substantial citations given the venues where they were published (32). Previous work on these extremely publishing non-research authors has questioned whether having non-experts with such a heavy presence and influence in the most visible literature is desirable (32). There is also evidence for advocacy bias affecting the COVID-19 literature where advocate authors with specific, biased viewpoints were given priority to spread massively their opinions (33,34).

Some limitations of our work should be discussed. First, our search strategy may have missed some relevant papers and may have had imperfect specificity. However, the voluminous nature of the COVID-19 literature is unquestionable. If anything, many journals are not indexed in Scopus, so the number of COVID-19 papers and authors may be an underestimate. Furthermore, our search focused on title-abstract-keywords, which miss many papers that may have secondary but still substantive mentions of COVID-19 material. Second, recall and precision for author IDs in Scopus is very high (19), but some authors may have been split in more than one ID profile, and some papers of different authors may cluster in the same ID profile. Some author ID profiles with 1 or a few papers are difficult to prove whether they are indeed different authors or simply fragments from the track record of a more prolific other author. The data on authors with at least 5 papers may thus be more reliable.

Acknowledging these caveats, our analysis demonstrates massive covidization and rapid ongoing decovidization of the scientific literature. While the COVID-19 field still has several important questions to address, including long-term outcomes and consequences of the pandemic and the pandemic response measures and interventions, a more targeted evolution of this literature and rational, focused efforts rather than diffuse over-productivity may be preferable.

## Data Availability

All key data are in the manuscript and supplements. Additional more detailed data may be requested from the authors.

---

**Supplementary Table 1.**
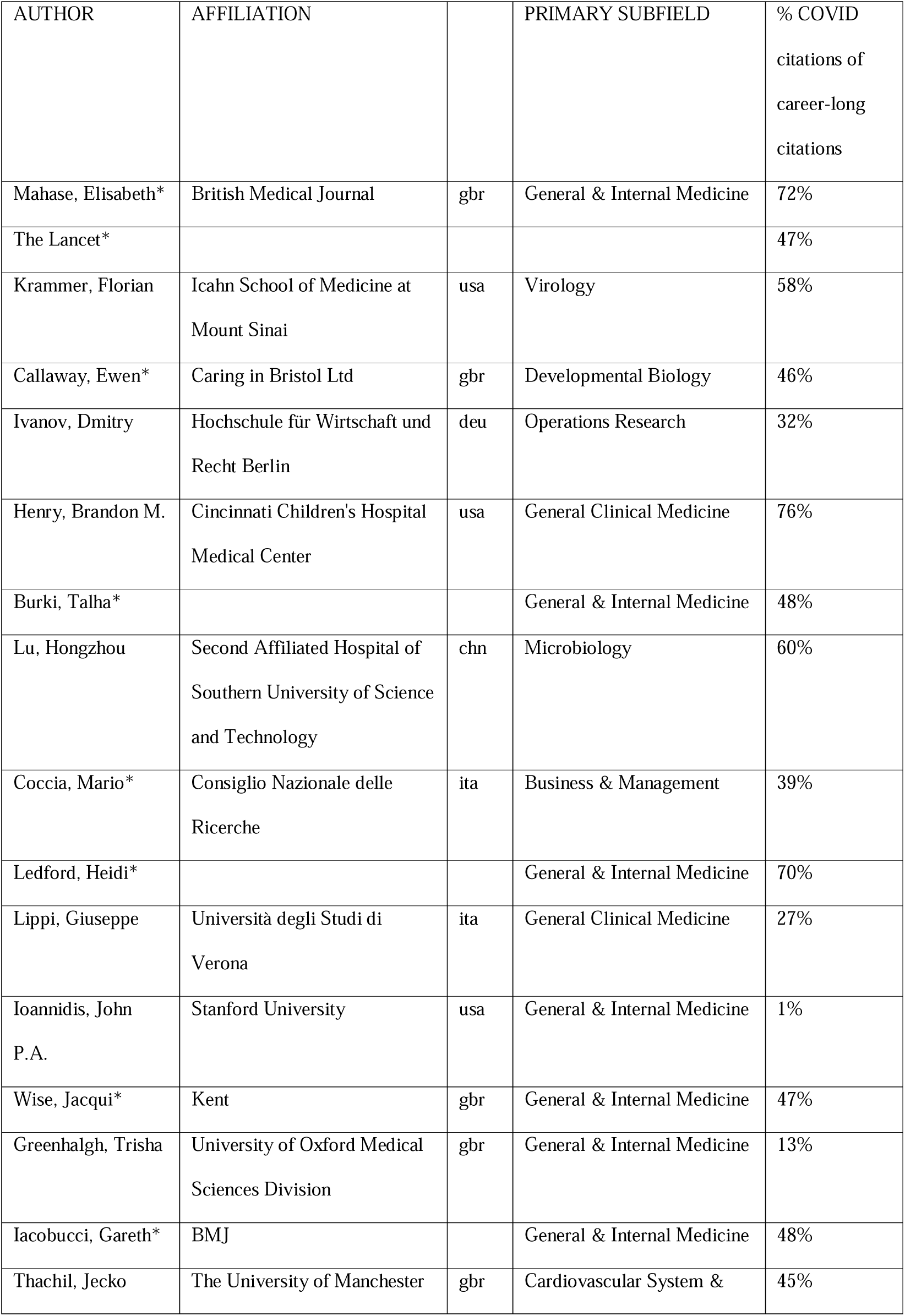
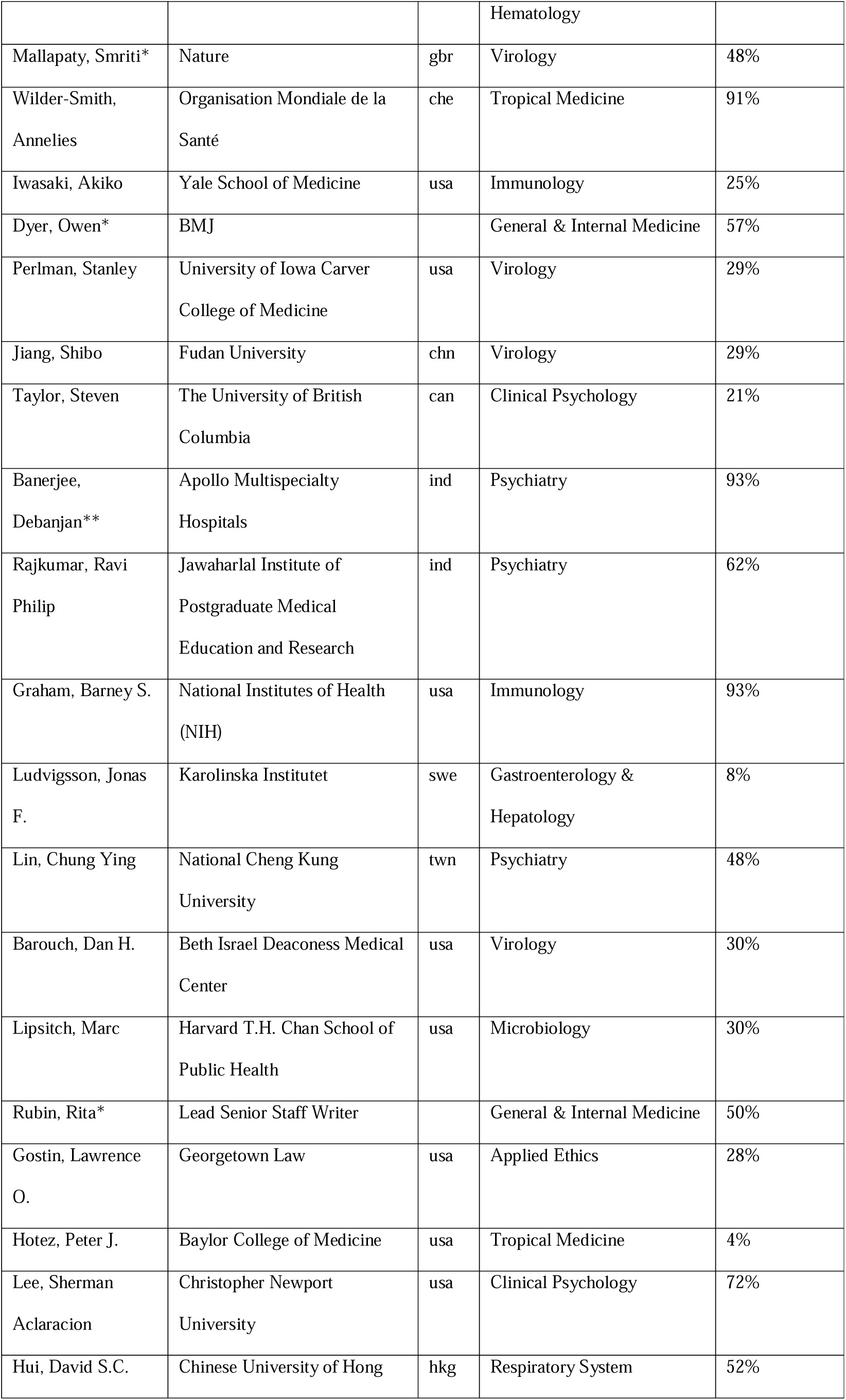
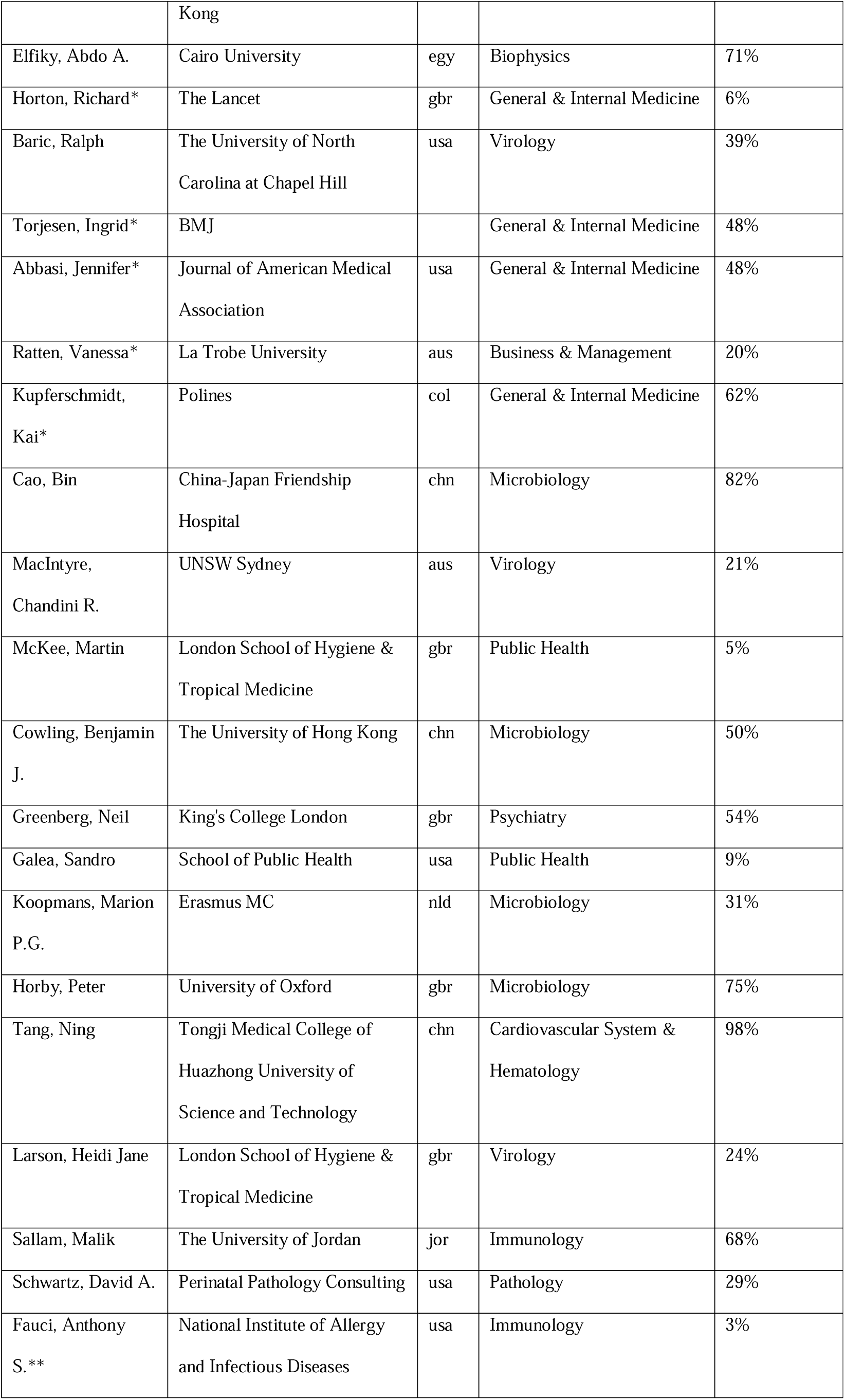
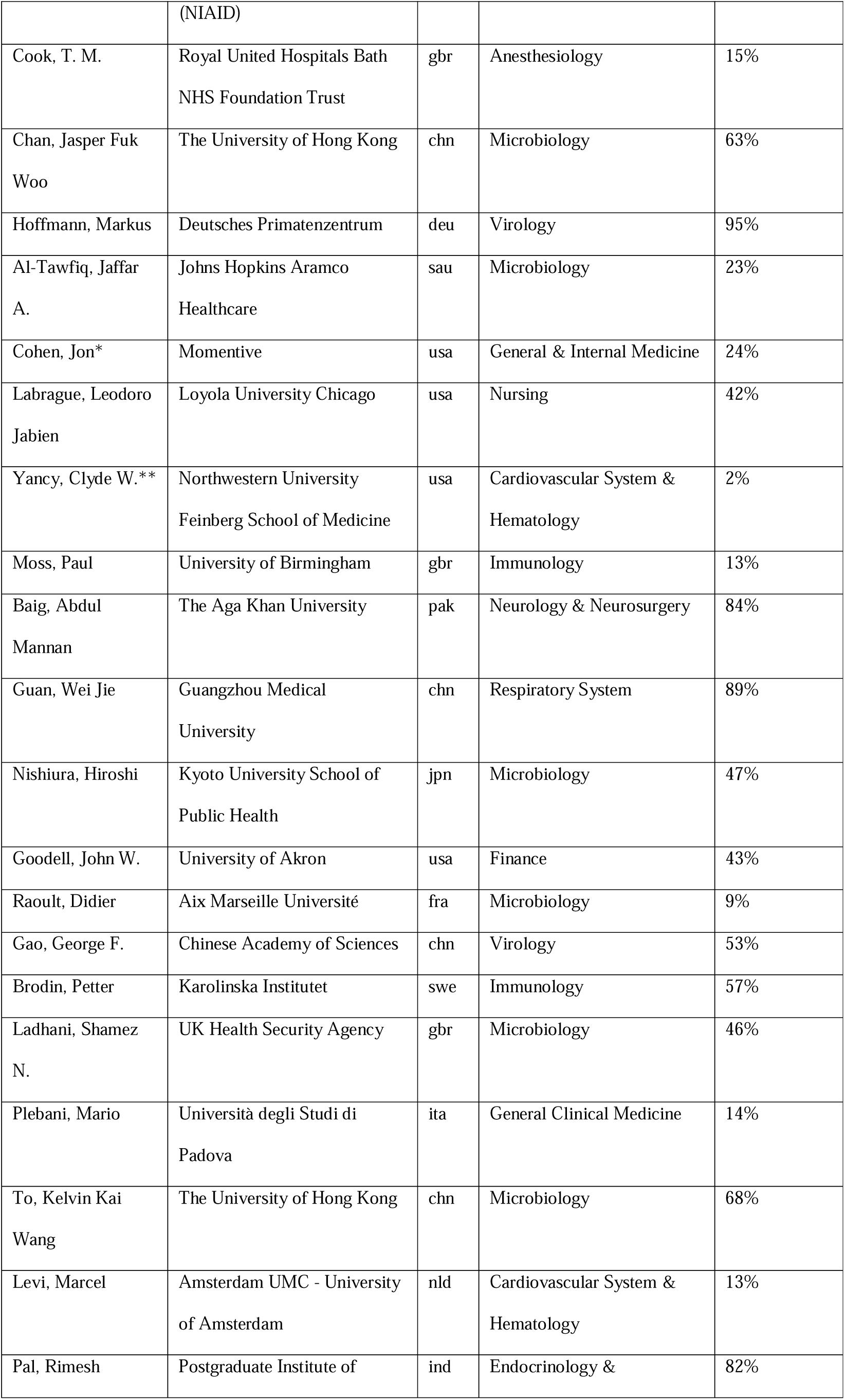
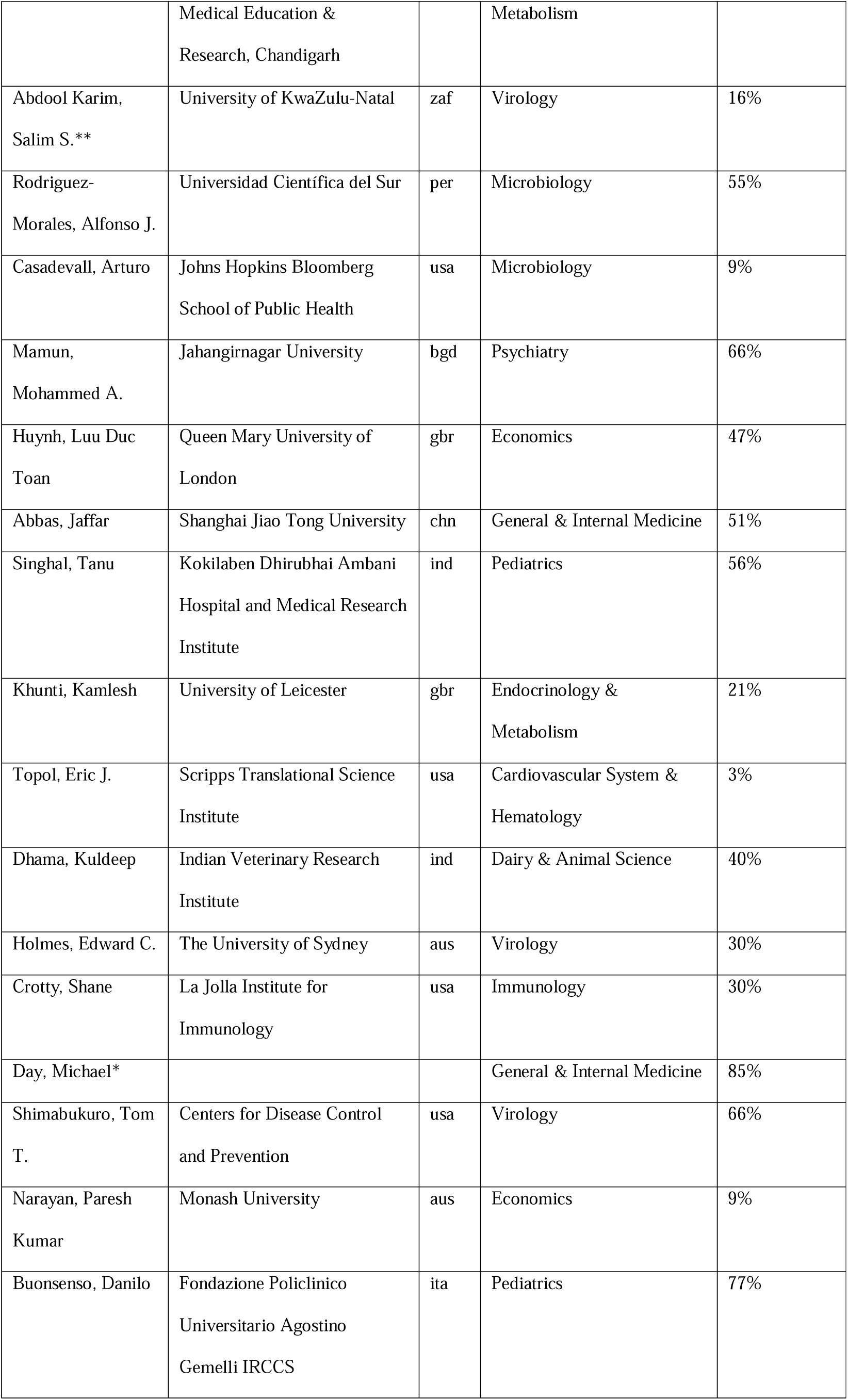
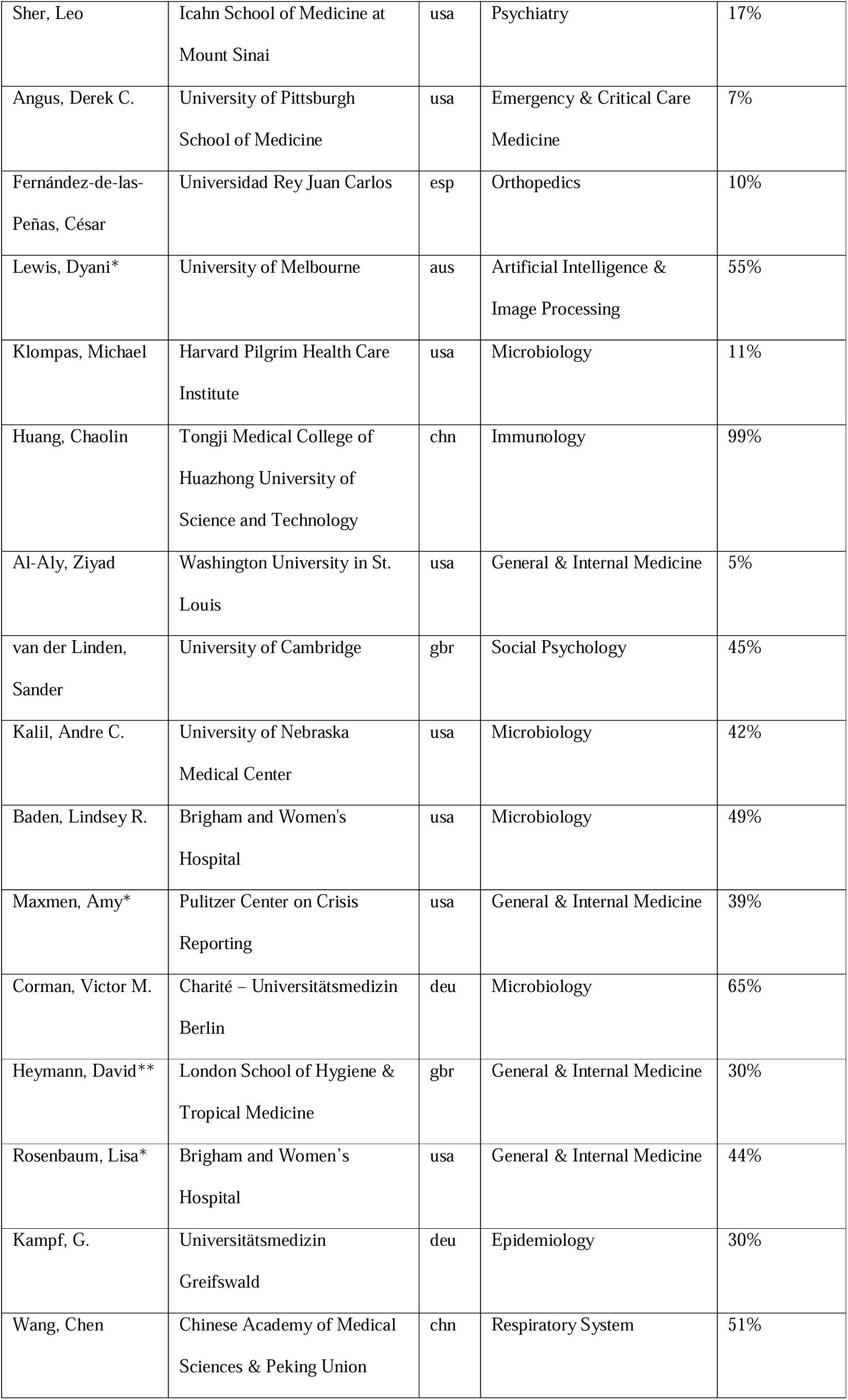
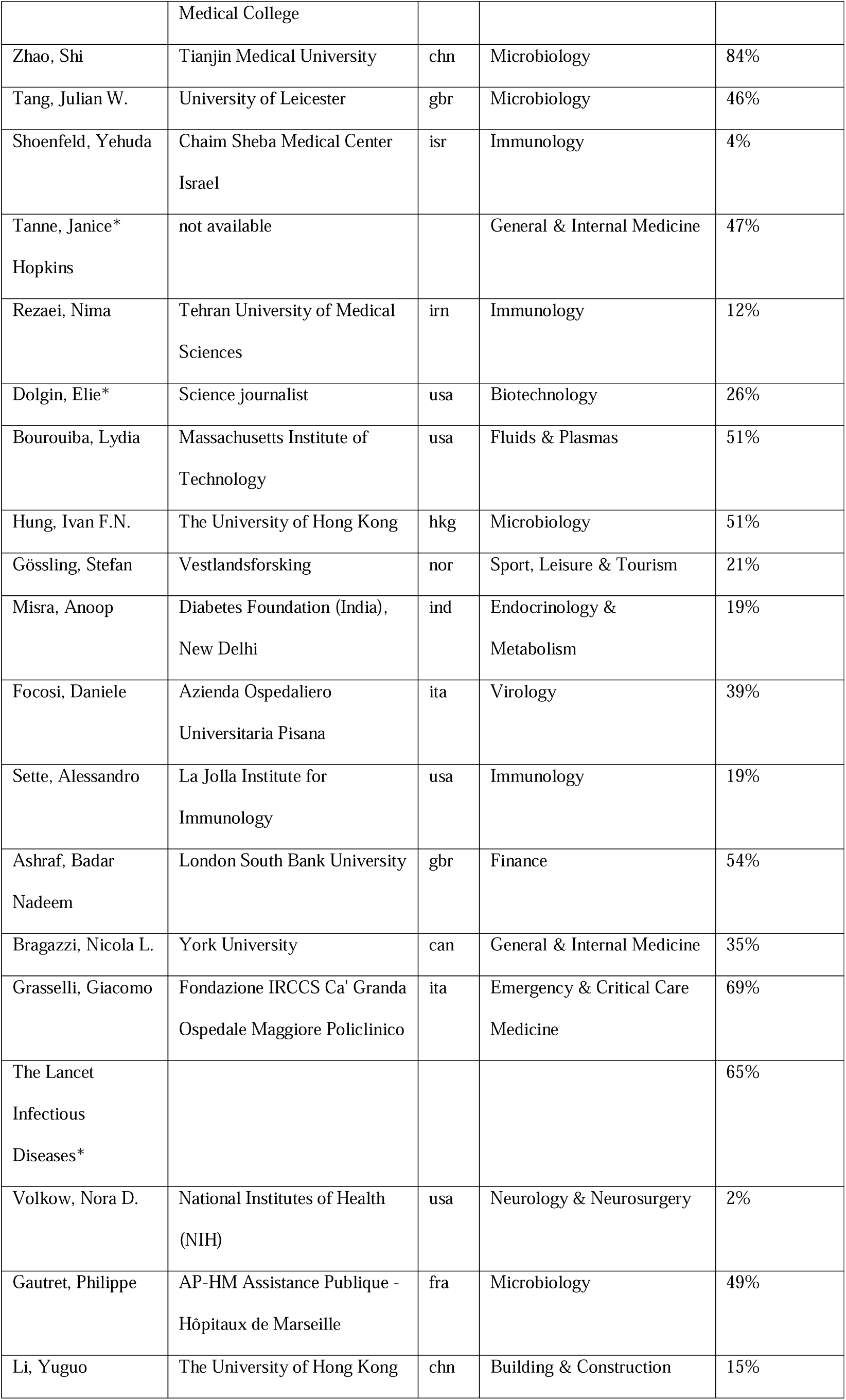
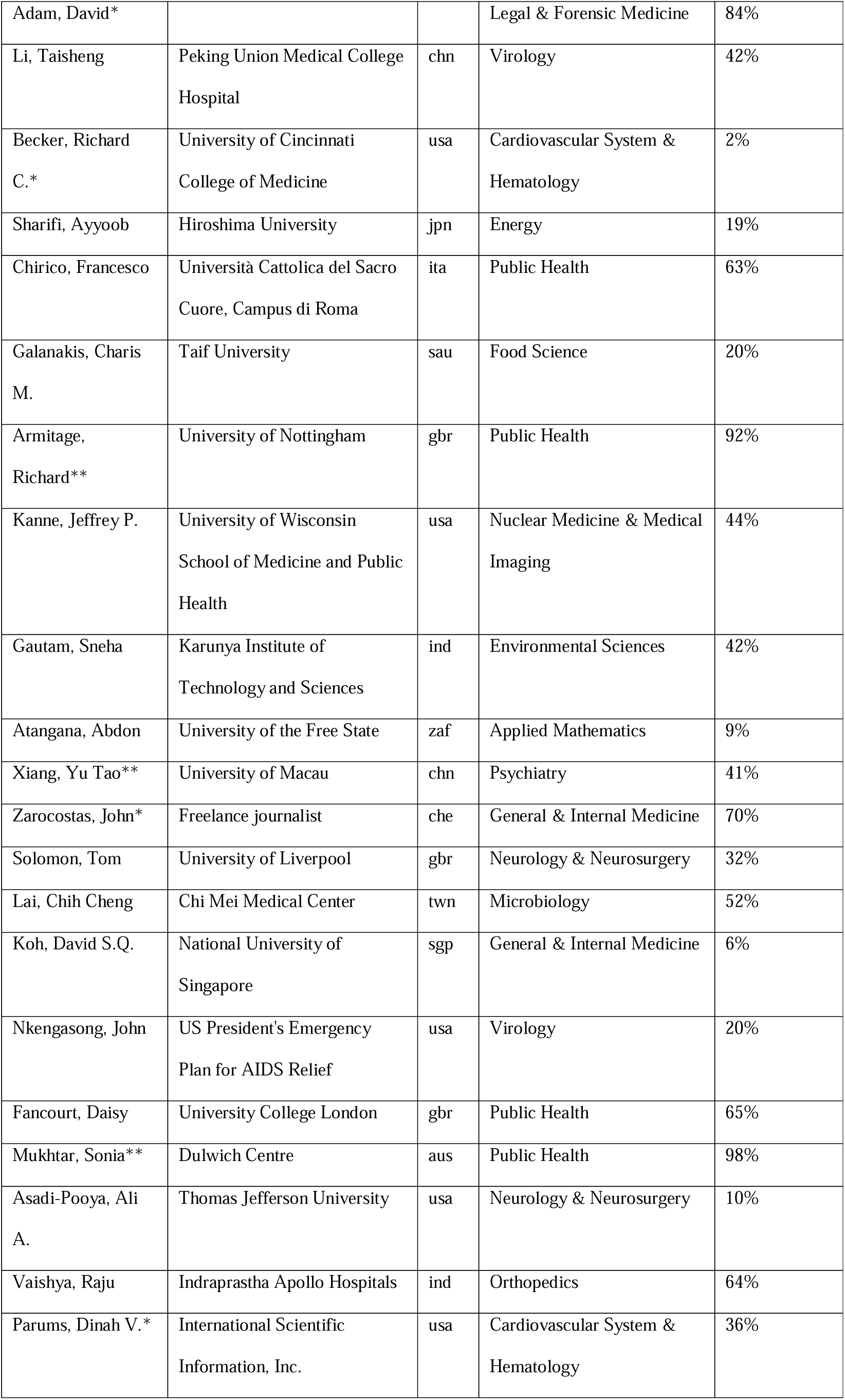
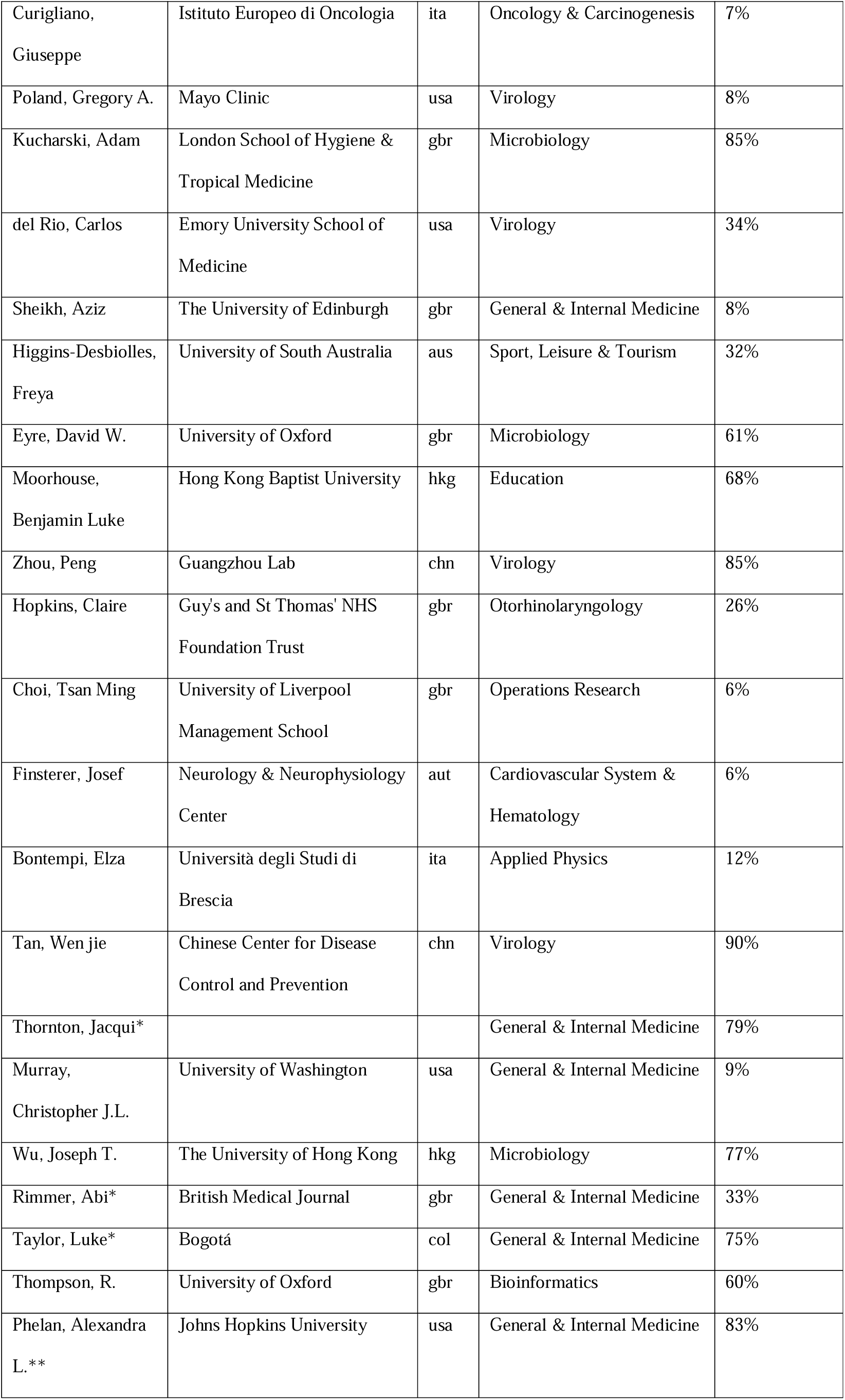
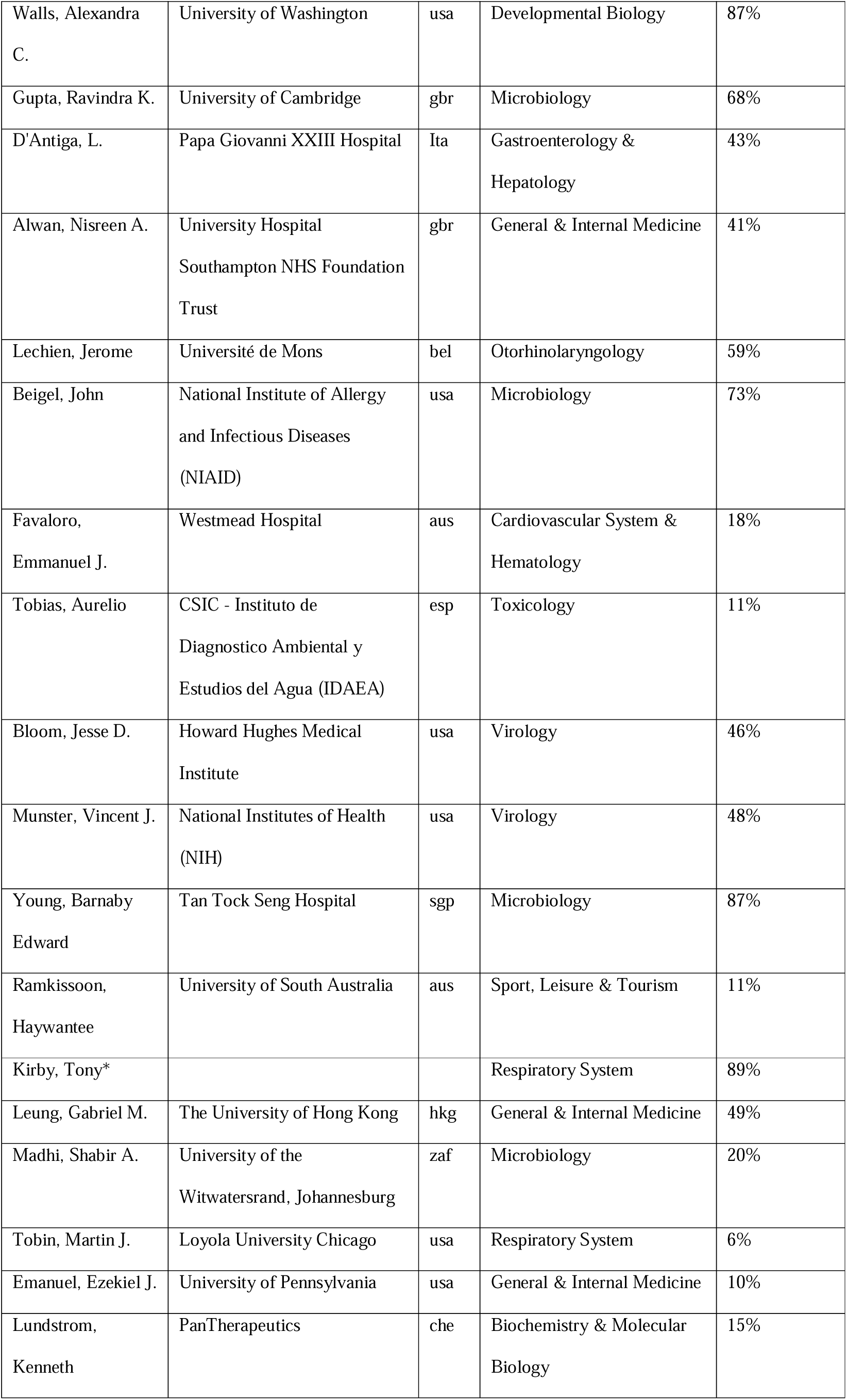
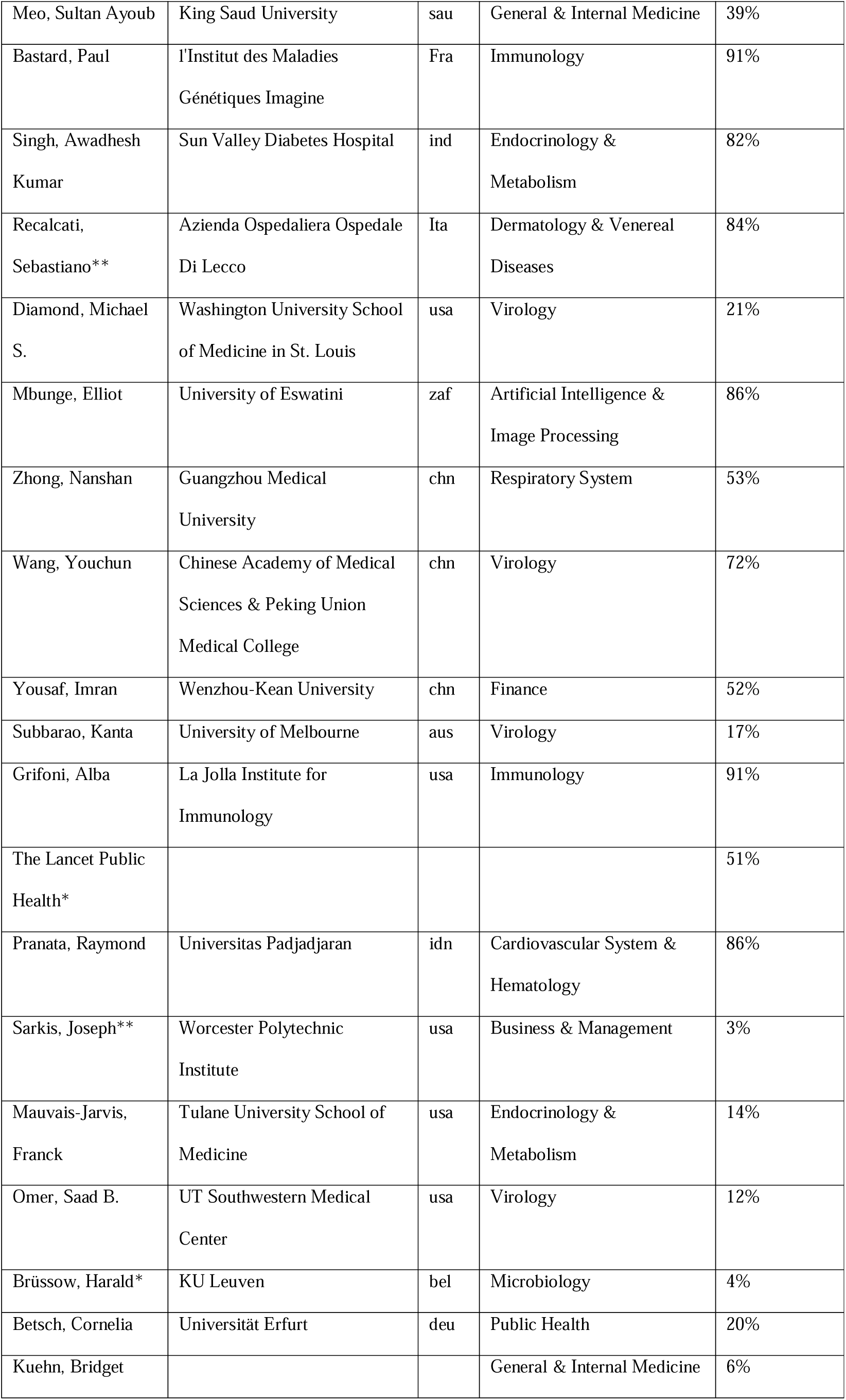
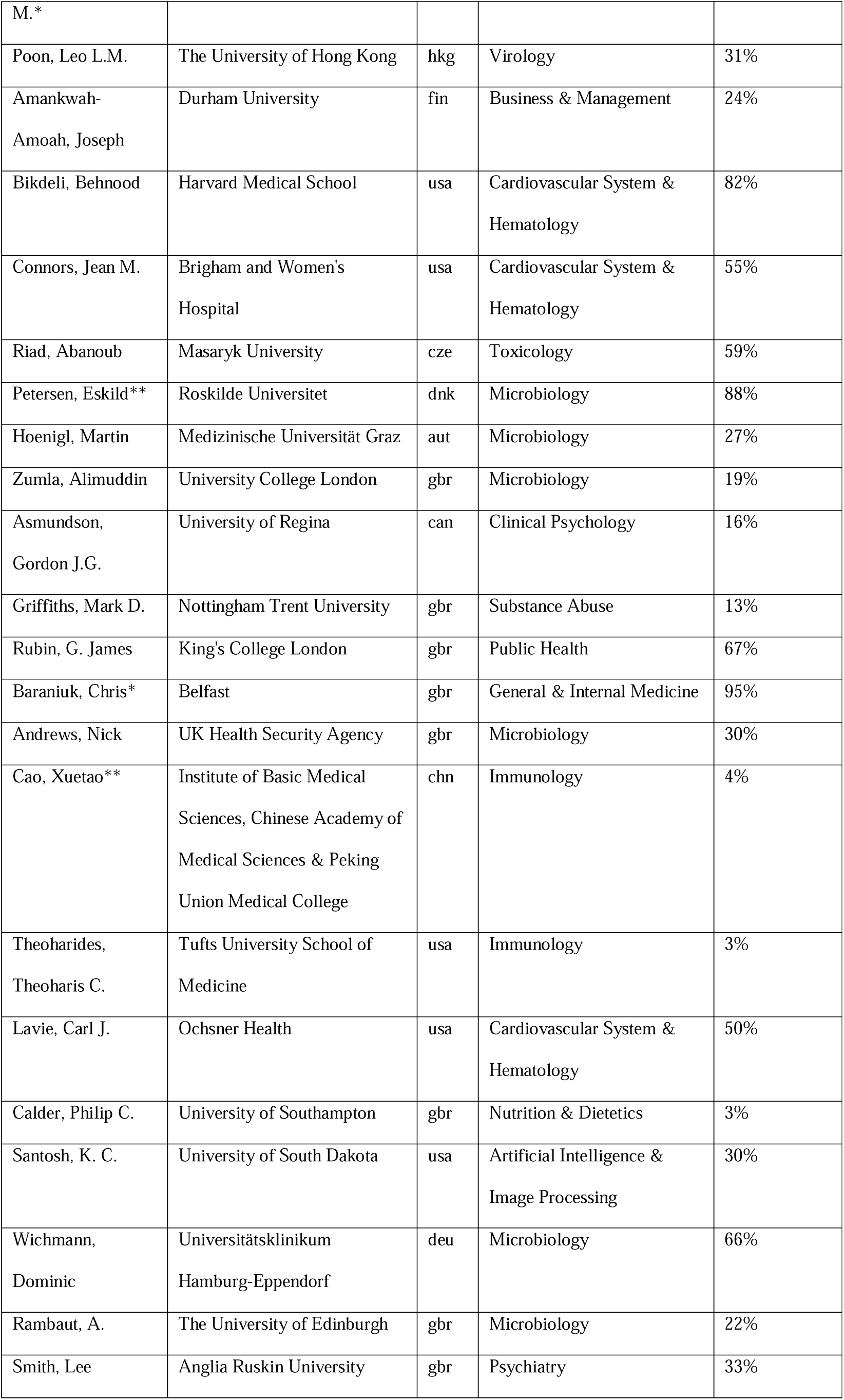
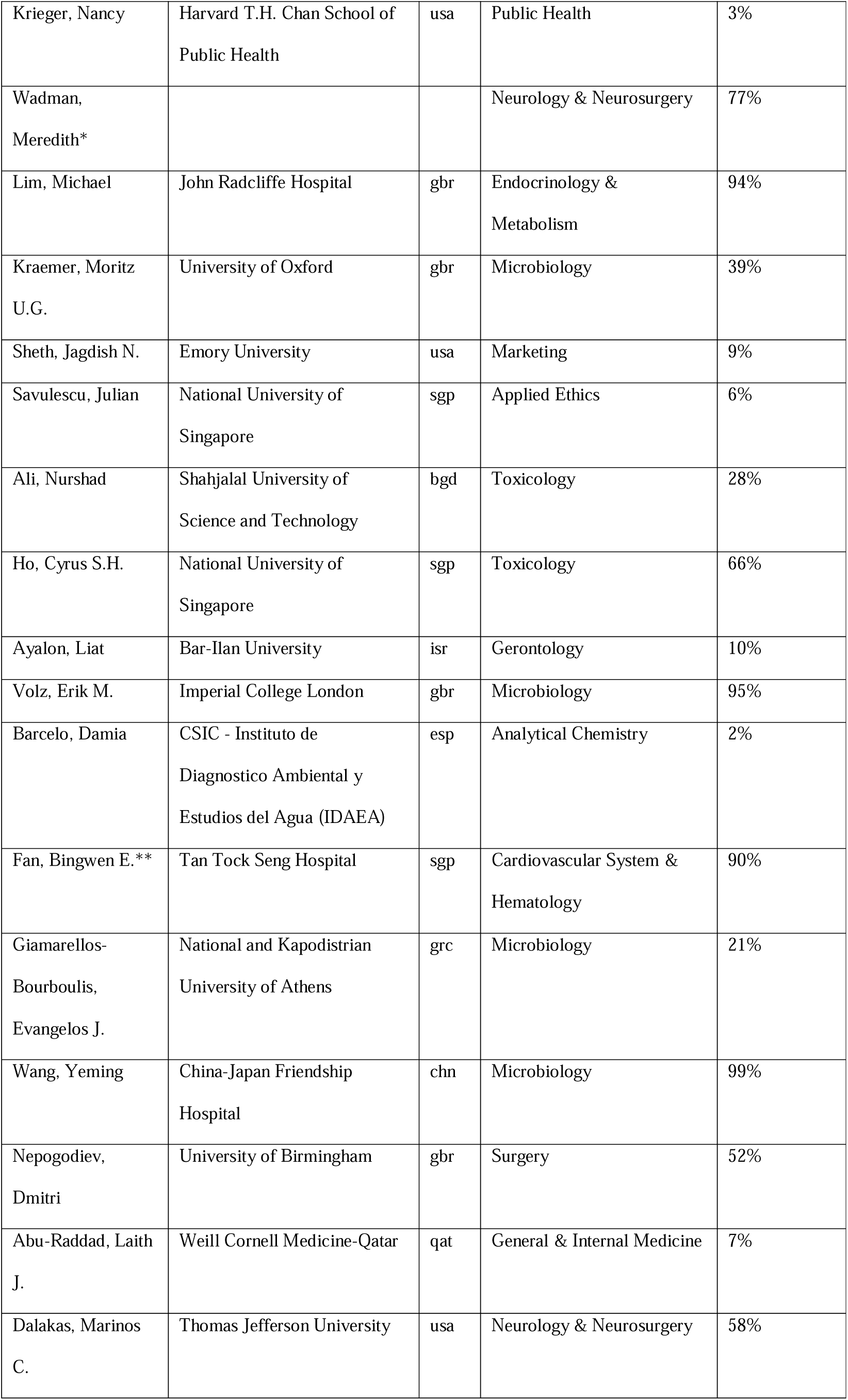
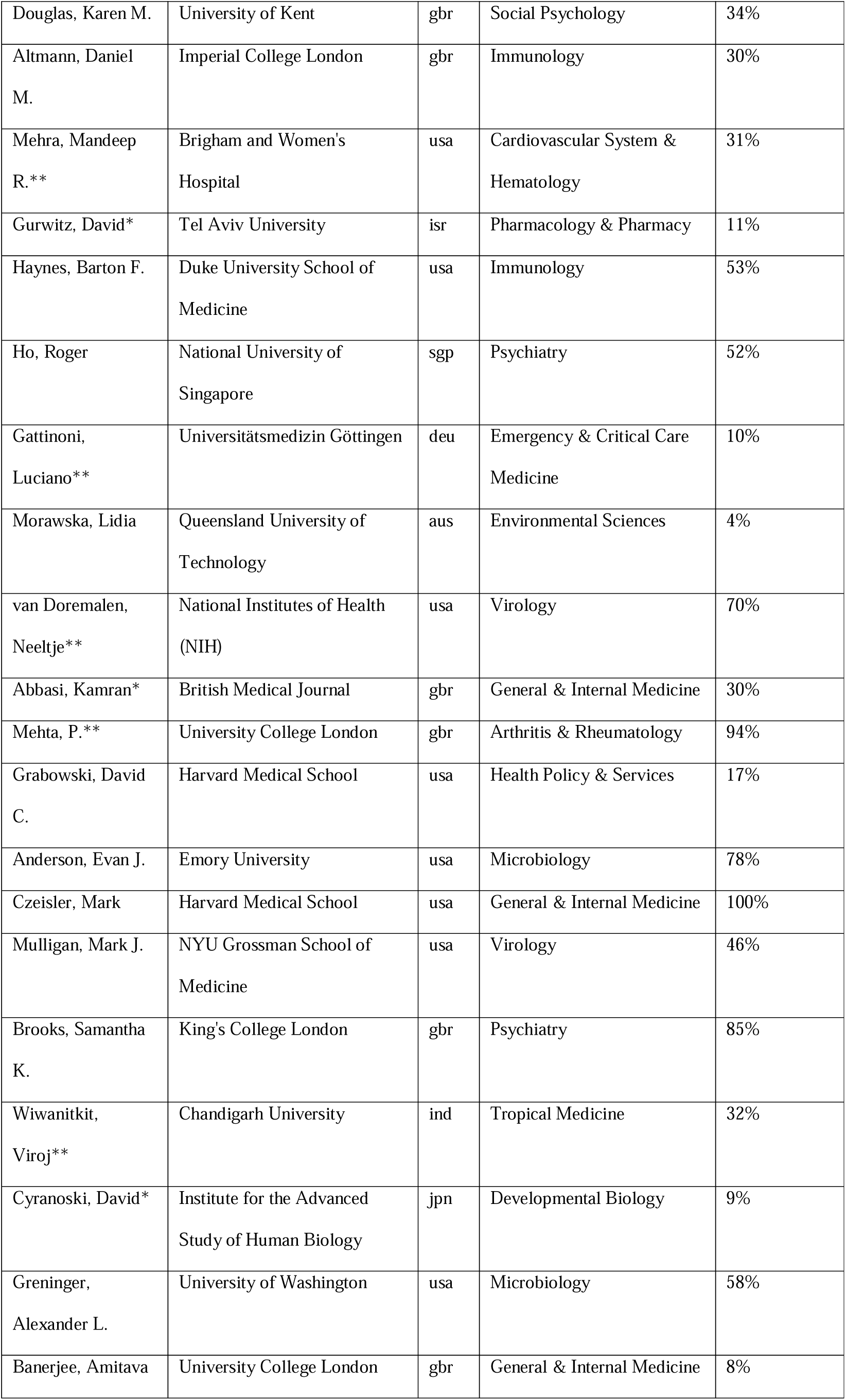
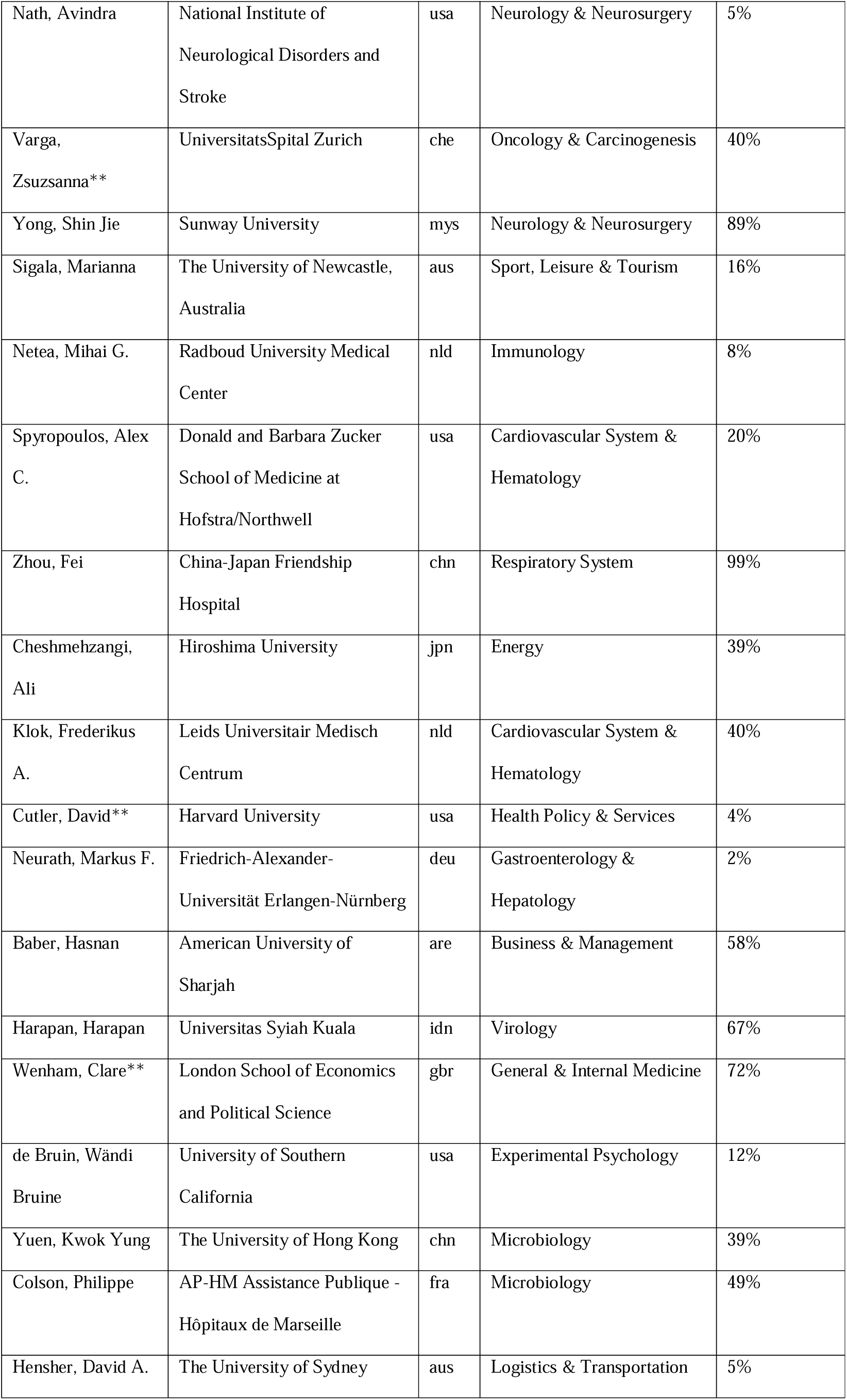
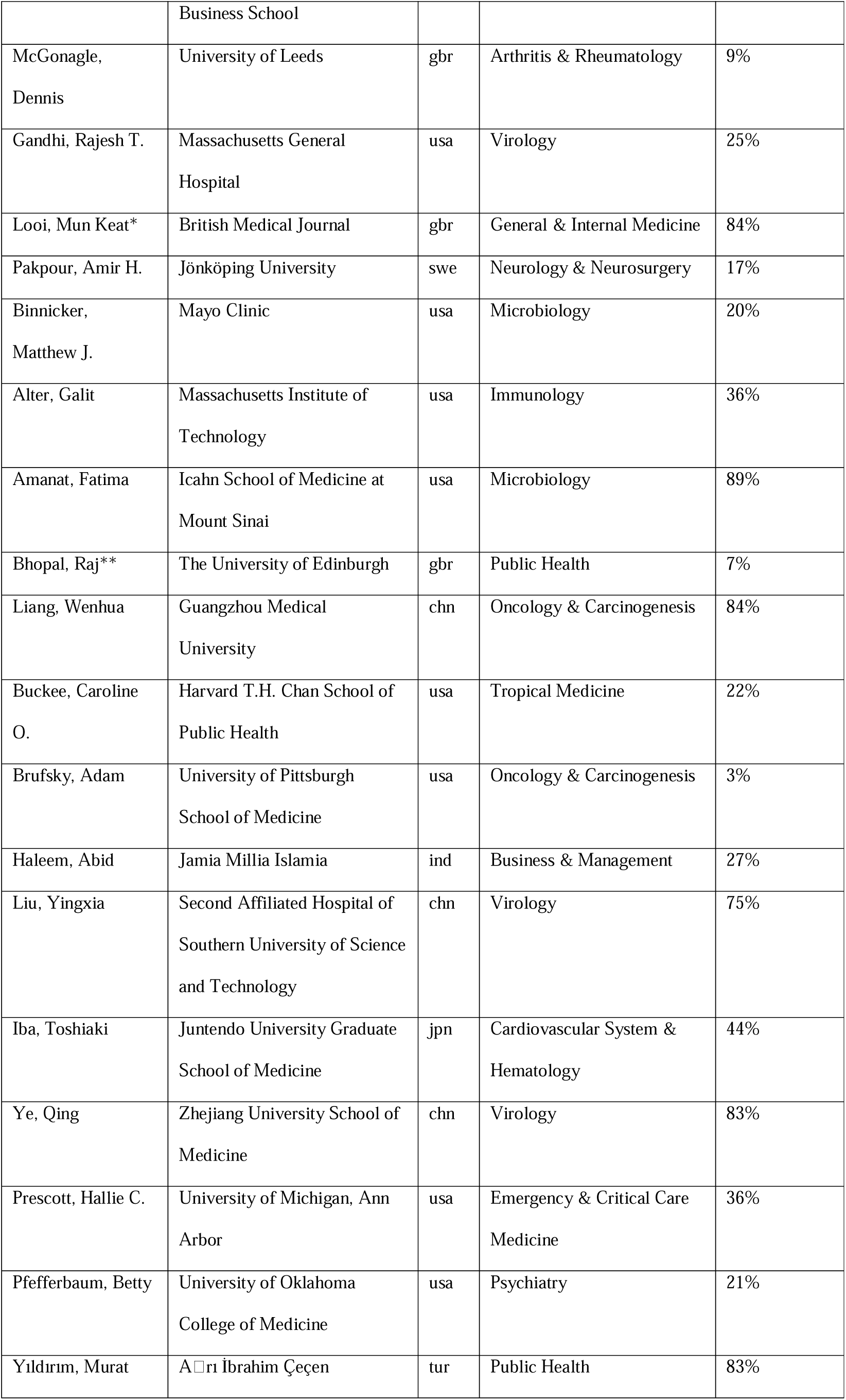
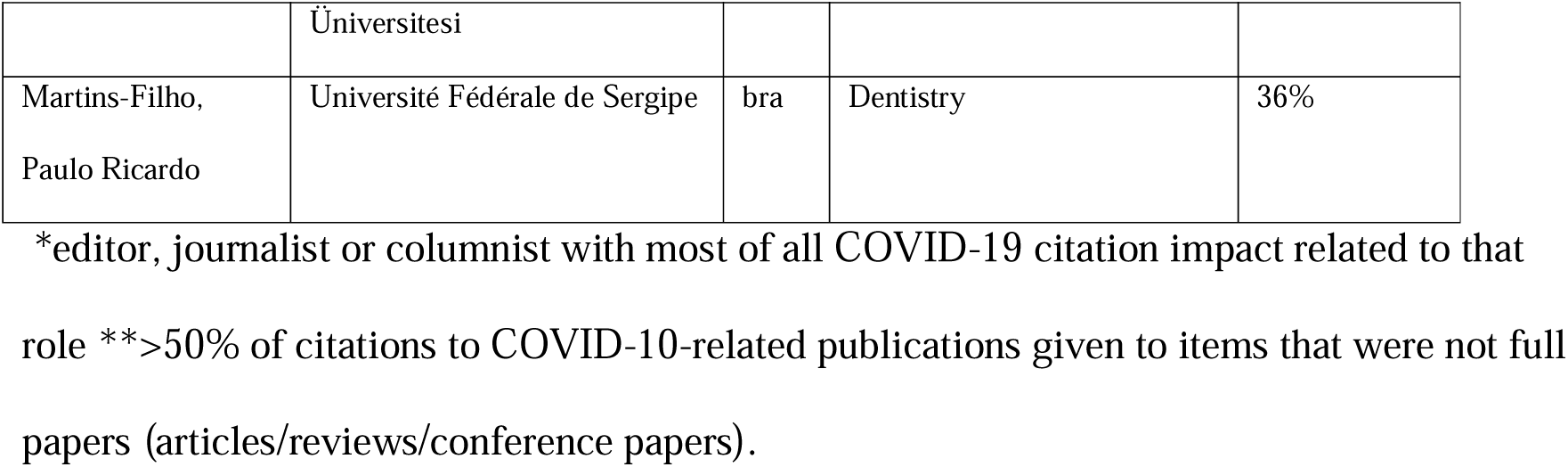
Top-300 authors with highest composite citation indicator for COVID-19-related published items during 2020-2024, excluding self-citations.

## REFERENCES

1. Ioannidis JPA, Salholz-Hillel M, Boyack KW, Baas J. The rapid, massive growth of COVID-19 authors in the scientific literature. R Soc Open Sci. 2021;8(9):210389.

2. Ioannidis JPA, Baas J, Klavans R, Boyack KW. A standardized citation metrics author database annotated for scientific field. PLoS Biol. 2019;17(8):e3000384.

3. Ioannidis JPA, Bendavid E, Salholz-Hillel M, Boyack KW, Baas J. Massive covidization of research citations and the citation elite. Proc Natl Acad Sci U S A. 2022;119(28):e2204074119.

4. Delardas O, Giannos P. How COVID-19 Affected the Journal Impact Factor of High Impact Medical Journals: Bibliometric Analysis. J Med Internet Res. 2022;24(12):e43089.

5. Pai M. Covidization of research: what are the risks? Nat. Med. 2020;26:1159.

6. Adam D. Scientists fear that ‘covidization’ is distorting research. Nature 2020;588:381–382.

7. Ioannidis JPA. The end of the COVID-19 pandemic. Eur J Clin Invest. 2022;52(6):e13782.

8. Delwiche FA. Letters to the editor on the Zika virus: a bibliometric analysis. J Med Libr Assoc. 2021;109(2):301–310.

9. Maalouf FT, Mdawar B, Meho LI, Akl EA. Mental health research in response to the COVID-19, Ebola, and H1N1 outbreaks: A comparative bibliometric analysis. J Psychiatr Res. 2021 Jan;132:198–206.

10. Kawuki J, Yu X, Musa TH. Bibliometric Analysis of Ebola Research Indexed in Web of Science and Scopus (2010-2020). Biomed Res Int. 2020 Sep 3;2020:5476567.

11. Zhang L, Zhao W, Sun B, Huang Y, Glänzel W. How scientific research reacts to international public health emergencies: a global analysis of response patterns. Scientometrics. 2020;124(1):747–773.

12. Oliveira JF, Pescarini JM, Rodrigues MS, Almeida BA, Henriques CMP, Gouveia FC, Rabello ET, Matta GC, Barreto ML, Sampaio RB. The global scientific research response to the public health emergency of Zika virus infection. PLoS One. 2020;15(3):e0229790.

13. Liang F, Guan P, Wu W, Liu J, Zhang N, Zhou BS, Huang DS. A review of documents prepared by international organizations about influenza pandemics, including the 2009 pandemic: a bibliometric analysis. BMC Infect Dis. 2018;18(1):383.

14. Hagel C, Weidemann F, Gauch S, Edwards S, Tinnemann P. Analysing published global Ebola Virus Disease research using social network analysis. PLoS Negl Trop Dis. 2017;11(10):e0005747.

15. Fricke R, Uibel S, Klingelhoefer D, Groneberg DA. Influenza: a scientometric and density-equalizing analysis. BMC Infect Dis. 2013;13:454.

16. Baas J, Schotten M, Plume A, Côté G, Karimi R. Scopus as a curated, high-quality bibliometric data source for academic research in quantitative science studies. Quant. Sci. Stud. 2020;1:377–386.

17. Baas J, Fennel C. When peer reviewers go rogue—estimated prevalence of citation manipulation by reviewers based on the citation patterns of 69000 reviewers. In Proc. of the 17th Int. Conf. of the Int. Soc. of Scientometrics and Informetrics (ISSI), Rome, Italy, 2019, pp. 963.–.

18. Archambault É, Beauchesne OH, Caruso J. 2011. Towards a multilingual, comprehensive and open scientific journal ontology. In Proc. of the 13th Int. Conf. of the Int. Soc. for Scientometrics and Informetrics (ISSI), Durban, South Africa, pp. 66–77.

19. Baas J, Boyack K, Ioannidis JPA. Data for updated science-wide author databases of standardized citation indicators. 2024, Doi: 10.17632/btchxktzyw.7)

20. Ioannidis JP, Klavans R, Boyack KW. 2016. Multiple citation indicators and their composite across scientific disciplines. PLoS Biol. 14, e1002501.

21. Ioannidis JPA, Boyack KW, Baas J. Updated science-wide author databases of standardized citation indicators. PLoS Biol. 2020;18: e3000918.

22. Maillard A, Delory T. Blockbuster effect of COVID-19 on the impact factor of infectious disease journals. Clin Microbiol Infect. 2022 Dec;28(12):1536–1538.

23. He J, Liu X, Lu X, Zhong M, Jia C, Lucero-Prisno DE 3rd, Ma ZF, Li H. The impact of COVID-19 on global health journals: an analysis of impact factor and publication trends. BMJ Glob Health. 2023 Apr;8(4):e011514.

24. Khatter A, Naughton M, Dambha-Miller H, Redmond P. Is rapid scientific publication also high quality? Bibliometric analysis of highly disseminated COVID-19 research papers. Learn Publ.2021; 34:568–577.

25. Wang D, et al., Abstracts for reports of randomised trials of COVID-19 interventions had low quality and high spin. J Clin Epidemiol 2021;139:107–120.

26. Li Y, et al., Reporting and methodological quality of COVID-19 systematic reviews needs to be improved: An evidence mapping. J Clin Epidemiol. 2021;135,:17–28.

27. Quinn TJ, et al. Following the science? Comparison of methodological and reporting quality of covid-19 and other research from the first wave of the pandemic. BMC Med. 2021;19:46.

28. Luo X, et al., Consistency of recommendations and methodological quality of guidelines for the diagnosis and treatment of COVID-19. J. Evid. Based Med. 2021;14:40–55.

29. Yang S, et al., Quality of early evidence on the pathogenesis, diagnosis, prognosis and treatment of COVID-19. BMJ Evid. Based Med. 2021;26:302–306.

30. Nieto I, Navas JF, Vázquez C. The quality of research on mental health related to the COVID-19 pandemic: A note of caution after a systematic review. Brain Behav Immun Health 2020;7:100123.

31. Chu JSG, Evans JA. Slowed canonical progress in large fields of science. Proc Natl Acad Sci U S A. 2021 Oct 12;118(41):e2021636118.

32. Ioannidis JPA. Prolific non-research authors in high impact scientific journals: meta-research study. Scientometrics. 2023;128(5):3171–3184.

33. Kepp KP, Cristea IA, Muka T, Ioannidis JPA. COVID-19 advocacy bias in the BMJ: meta-research evaluation. medRxiv 2024.06.12.24308823; doi: 10.1101/2024.06.12.24308823.

34. Kepp KP, Aavitsland P, Ballin M, Balloux F, Baral S, Bardosh K, Bauchner H, Bendavid E, Bhopal R, Blumstein DT, Boffetta P, Bourgeois F, Brufsky A, Collignon PJ, Cripps S, Cristea IA, Curtis N, Djulbegovic B, Faude O, Flacco ME, Guyatt GH, Hajishengallis G, Hemkens LG, Hoffmann T, Joffe AR, Klassen TP, Koletsi D, Kontoyiannis DP, Kuhl E, La Vecchia C, Lallukka T, Lambris J, Levitt M, Makridakis S, Maltezou HC, Manzoli L, Marusic A, Mavragani C, Moher D, Mol BW, Muka T, Naudet F, Noble PW, Nordström A, Nordström P, Pandis N, Papatheodorou S, Patel CJ, Petersen I, Pilz S, Plesnila N, Ponsonby AL, Rivas MA, Saltelli A, Schabus M, Schippers MC, Schünemann H, Solmi M, Stang A, Streeck H, Sturmberg JP, Thabane L, Thombs BD, Tsakris A, Wood SN, Ioannidis JPA. Panel stacking is a threat to consensus statement validity. J Clin Epidemiol. 2024;173:111428.

